# The roles of pre-season immunity, age, viral shedding, and community exposures in shaping influenza household transmission dynamics

**DOI:** 10.1101/2025.03.25.25324622

**Authors:** Molly K Sauter, Jackie Kleynhans, Jocelyn Moyes, Meredith L McMorrow, Florette K Treurnicht, Orienka Hellferscee, Anne von Gottberg, Nicole Wolter, Amelia Buys, Lorens Maake, Neil A Martinson, Kathleen Kahn, Limakatso Lebina, Katlego Motlhaoleng, Floidy Wafawanaka, Francesc Xavier Gómez-Olivé, Stefano Tempia, Bryan Grenfell, Cecile Viboud, Kaiyuan Sun, Cheryl Cohen

## Abstract

Our understanding of influenza transmission remains imperfect due to the high prevalence of asymptomatic infections that often go undetected. To address this challenge, we leveraged uniquely resolved data from a household cohort study spanning three consecutive years in rural and urban South Africa. The study incorporated pre-season serum collection and twice-weekly virological testing during the influenza season, regardless of symptom presence. We developed a subtype/lineage-specific influenza household transmission model that accounts for time-resolved viral shedding across the full clinical spectrum of infections, allowing us to disentangle the role of household and community exposures, pre-season immunity, and age on transmission. Our analysis revealed that viral shedding intensity, as measured by the cycle threshold (Ct) values of infected household members, significantly correlated with the risk of transmission for all four influenza subtypes/lineages. After adjusting for viral shedding, pre-season hemagglutination inhibition (HAI) titers greater than 1:40 were associated with a significantly lower risk of infection acquisition for A(H1N1)pdm09, A(H3N2), and B/Victoria, but not for B/Yamagata. Notably, children exhibited higher susceptibility and longer viral shedding durations compared to adults across all subtypes/lineages, even after adjusting for pre-season HAI titers. While our findings support that HAI titers correlate with protection, the strong residual effects of age on susceptibility and viral shedding may reflect the accumulation of additional immune responses shaped by repeated exposures over time. Our study underscores the need to explore immune mechanisms beyond HAI titers that modulate influenza susceptibility and transmission.

## Introduction

Refining our understanding of influenza transmission is essential for identifying key risk factors for infection, characterizing protective immunity, and informing prevention strategies to reduce disease burden. Human challenge studies have provided valuable insights into the role of prior immunity and exposure dose on infection risk. Yet, these studies involve healthy volunteers in controlled laboratory settings and may not adequately represent the diverse populations and real-world contexts in which influenza spreads ^1,2^. In parallel, studying influenza transmission in real-world settings is challenging due to the acute nature of infection, with viral shedding typically lasting only 3 to 6 days, and a high prevalence of asymptomatic infections ^1,3–5^. Consequently, the two observational study designs traditionally used to assess influenza transmission and protective immunity – household cohort studies and case-ascertained household transmission studies – often encounter difficulties in tracking infections and capturing transmission dynamics ^6^. Overcoming these challenges through improved study designs remains a priority to enhance our understanding of influenza transmission and inform more effective intervention strategies.

The Prospective Household cohort study of Influenza, Respiratory Syncytial virus, and other respiratory pathogens community burden and Transmission dynamics in South Africa (PHIRST) aims to address these challenges by introducing the collection of pre-season serum samples followed by twice-weekly respiratory virological testing regardless of symptom presence throughout the influenza season ^7^. This unique design leverages the distinct strengths of traditional observational studies while overcoming key limitations. Prior household cohort studies have sampled based on symptomatic illness and/or inferred infections from paired sera. Although the paired sera approach can detect asymptomatic and subclinical infections, its sensitivity and specificity are limited, and it lacks precision in determining the timing of infection ^6,8–10^. Case-ascertained household transmission studies have relied on clinical visits of symptomatically infected individuals to enroll index cases and their households, neglecting transmission events initiated by asymptomatic individuals and those who do not seek care ^5,6,11,12^. In contrast, systematic and asymptomatic testing in the PHIRST cohort allows for the precise identification of influenza infections and household index cases across the clinical spectrum of disease, eliminating the bias towards symptomatic transmission. Further, the PHIRST cohort enables the assessment of the relationship between pre-existing immunity and the risk of infection ^6,8–10^, along with direct observation of transmission events through repeated virological testing at short time intervals ^5,6,11^. Specifically, these data provide the means to reconstruct viral shedding kinetics for each influenza-infected participant during the study period, which permits the characterization of time-varying influenza exposure risk for susceptible household members.

In this study, leveraging the PHIRST study’s unique design and comprehensive data collection in a high transmission setting, we developed a highly detailed household transmission model to understand how influenza virus exposure intensity, age, and serum HAI titers influence the risk of infection acquisition. We are particularly interested in quantifying the role of prior immunity while accounting for influenza virus exposure, and vice versa. We also provide valuable insights into influenza virus transmission from low- and middle-income countries, where the burden of influenza is heightened by low vaccination rates, population demographics, and a high prevalence of comorbidities ^13–16^.

## Methods

### Study design

The PHIRST study enrolled participants from two study sites in rural and urban communities of South Africa^7^. The rural site was in the Agincourt sub-district of the Mpumalanga Province Demographic Surveillance Site, an established health and socio-demographic surveillance site (HDSS) since 1992. The urban study site was the Jouberton suburb of Klerksdorp in the North West Province. New cohorts of households were enrolled each year from 2016 to 2018 prior to the start of the winter influenza season, which typically runs from May to September in South Africa’s temperate climate. In Agincourt, households were randomly selected from the HDSS database and in Klerksdorp, selection was based on proximity to randomly generated global positioning system (GPS) coordinates. Successful enrollment required that the household had more than 2 members and that at least 80% of the members provided written consent, or written assent and consent from a parental guardian for household members <17 years old. Each year we enrolled an average of 55 new households, or 281 individuals, per study site. A total of 327 households and 1684 individuals were successfully enrolled and completed follow-up surveys and samples over the duration of the three-year study.

Upon enrollment we collected demographic, household, and baseline health data, including an offer of HIV testing to all participants who did not have a known HIV status and were over 10 years old; children were tested at birth and, when negative, were assumed to be negative through age 10. Blood serum samples were collected at least once prior to each influenza season. After enrollment the participants entered an intensive follow up period for six months in 2016, and 10 months in 2017 and 2018, spanning the entirety of the influenza season. Trained investigators visited each household twice weekly to collect nasopharyngeal swabs regardless of the presence of symptoms or health-seeking behavior.

### Laboratory Testing

Influenza antibody titers were measured with HAI assays against strains circulating during the study period: A/South Africa/2517/2016 (A(H1N1)pdm09), A/Singapore/INFIMH-16-0019/2016 (A(H3N2)), B/South Africa/R3037/2016 (B/Victoria), and B/South Africa/R05631/2017 (B/Yamagata). The same viruses were used for all three study years. Nasopharyngeal swabs were tested for influenza with rRT-PCR using the FTD Flu/RSV detection assay (Fast Track Diagnostics, Luxembourg) ^7,17^. Positive samples were further analyzed to determine the influenza subtype or lineage with the US CDC influenza A subtyping kit or the CDC influenza B Yamagata and Victoria lineages typing kit (available through International Reagent Resource Program; http://www.internationalreagentresource.org) ^7,17^. The cohort design, enrollment, testing, and laboratory protocols have all been described in detail in previous publications ^7^.

### Analytical Approach

To build a household transmission model that captures the daily risk of influenza virus infection based on the shedding dynamics of infected household members, we first estimated the viral shedding kinetics of each observed infection episode. Episodes were defined by clustering influenza-positive samples (rRT-PCR cycle threshold (Ct) value <37) that were less than 14 days apart and of the same subtype ^3^. Some samples tested positive for an influenza type but did not reach the threshold for positivity on the corresponding subtype or lineage assay. If non-subtyped samples were temporally clustered with samples that could be subtyped, they were assumed to be a part of the same infection episode. We observed limited instances of co-circulation of influenza virus subtypes causing 25 instances of coinfection (4% of all observed infections) which were excluded from our analysis, as it is challenging to attribute the Ct value accurately to a single subtype in the presence of co-infection.

The rRT-PCR Ct value has been shown to provide a quantitative proxy (though not a direct measure) of a sample’s viral load and has been used in previous studies of influenza transmission ^1,5,18^. Based on previous work, we assumed an exponential increase in viral load during the proliferation stage, defined as the period from the onset of shedding to the peak viral load, followed by an exponential decrease during the clearance stage, defined as the period from the peak viral load to the end of shedding ^19,20^. This translates to a linear decrease in Ct value from the threshold value of 37 to a minimum Ct value (i.e., proxy peak viral load), followed by a linear increase in Ct from the minimum to detection threshold. Hence, shedding kinetics can be modeled as a piecewise linear function and fitted to serial Ct values of each infection episode, where Ct values are based on the multiplex FTD Flu/RSV detection assay. To calibrate the model, we implemented a Bayesian hierarchical framework using the Hamiltonian Monte Carlo (HMC) algorithm to generate posterior draws for parameter inference. Each draw estimates the value and timing of the minimum Ct (a proxy for peak viral load), as well as the durations of the proliferation and clearance phases, both at the population mean for each subtype or lineage and at the individual (per-infection episode) level. From the individual model estimates and our knowledge of the timing of the infection episode within the season, we determined start and end dates of each infection episode to establish a timeline of infection within the household and study site. The onset of infection was set as one day prior to the first day of shedding based on kinetics observed in challenge studies ^1,2^ (for model equations and details see Supplementary Text). In all following regression analysis, to account for uncertainty in the shedding estimates due to potential censoring or missing observations, we resampled from the posterior; each regression was run 100 times, drawing one random posterior estimate per individual from the shedding model outputs, and the resulting regression estimates were pooled to obtain the final reported values.

Next, we explored individual factors that are predictive of viral shedding duration and peak viral load (estimated minimum Ct). We used a mixed effects multiple linear regression fit separately to each subtype/lineage and viral kinetics characteristic (total shedding duration and peak viral load) with fixed effects covariates including age, sex, and HIV infection status and year as a random effect; as B/Yamagata only circulated in 2017, a multiple linear regression with no random effect was used. HIV status was included as a covariate given the high prevalence of participants living with HIV (PLWH) in the study population and its potential relevance to influenza transmission dynamics. HAI antibody titers are commonly used serologic markers of protective humoral immunity to influenza infection with a titer cutoff of 1:40 described to provide 50% protection against the infection ^21–23^. Thus, we also included the participant’s pre-season HAI titer grouped as either above or equal to 1:40 or less than 1:40 as an indicator of pre-season immunity. For the majority of individuals, we used the geometric means of two draws from a serological sample taken closest to the start of the influenza season, between March and May; however, for the 87 (5.7%) individuals that did not have a blood draw in this time period, we interpolated their titer measurement based on the initial serological measurement taken between November and February. To do so, for individuals that had both a draw in November-February and March-May, we linearly regressed the March-May titer draw against the November-February draw as well as a selection of age, sex, and HIV status of the individual that created the best fit transmission model for each subtype (for model testing see Supplementary Tables 2 and 3). We then used this regression to interpolate the March-May draw based on the November-February draw for those missing this second draw. The shedding regression models were fit separately to each influenza subtype/lineage.

To assess predictors of influenza virus infection risk, we employed piecewise exponential survival models with time-varying covariates. These models integrated daily estimates of viral shedding intensity, community influenza prevalence, and other relevant covariates^20,24,25^. The piecewise exponential model was estimated using Poisson regression, with each individual’s daily infection status as the binary outcome and corresponding daily covariates. At the start of the season, all individuals were considered susceptible and were then excluded from the risk set following the onset of infection with a specific subtype or lineage. Although individuals were no longer deemed susceptible after infection onset, their infection status and shedding intensity continued to contribute to the transmission risk for other household members. A time-varying covariate representing the household force of infection (FOI) was calculated by summing the estimated daily shedding intensities of all influenza-infected individuals within the household. We also included a community FOI term, derived from the prevalence of each influenza subtype within the cohort at the corresponding study site and year. This was measured as the daily number of PCR-positive individuals divided by the total population at the site (see Supplementary Fig 10). Data for A(H1N1)pdm09 and B/Victoria included observations from both 2016 and 2018, while observations for A(H3N2) and B/Yamagata were restricted to 2017. Additional individual-level covariates included pre-season HAI titer, age, sex, HIV status, and household size. Further methodological details are provided in the Supplementary Text.

## Results

### Cohort Description

The PHIRST participants were young (56% of the individuals were <18 years old) and resided in large households (40% of households had over 5 members). HIV prevalence was high with 16% of participants living with HIV (PLWH) and 19% of PLWH having a CD4 count less than 200 (an indication of immune suppression). No individuals included in this analysis had received an influenza vaccine for the then-current year. Table 1 lists individual and household characteristics for individuals that completed the follow-up period, provided all necessary samples, and had pre-season titer samples analyzed (see ^7^ for a complete cohort profile). Over the three years of the study, there were 1258 influenza-positive specimens, comprising around 1% of all the specimens collected. Among the 1518 individuals included in the analysis, 513 (34%) experienced at least one influenza virus infection episode within the year of the individual’s study enrollment. All four known circulating subtypes/lineages of seasonal influenza were captured over the course of the three-year study. In 2016, the shortest data collection period, the predominant subtypes/lineages were A(H1N1)pdm09 and B/Victoria, though there was some co-circulation of A(H3N2). Conversely, A(H3N2) and B/Yamagata led circulation in 2017. A(H1N1)pdm09 and B/Victoria returned to predominance in 2018 (for sampling results across all years, see Supplementary Figs 4-6).

**Table 1.**
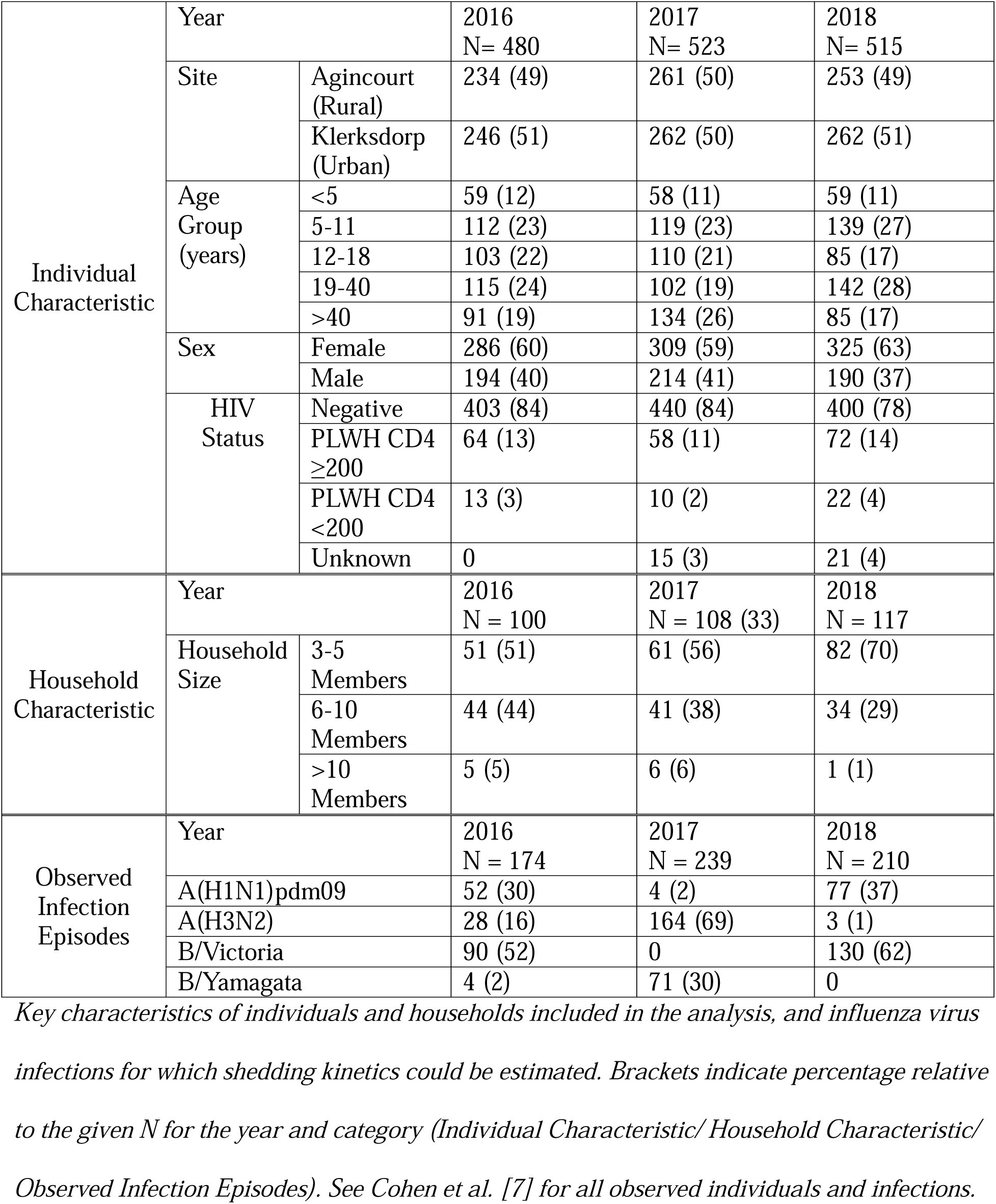
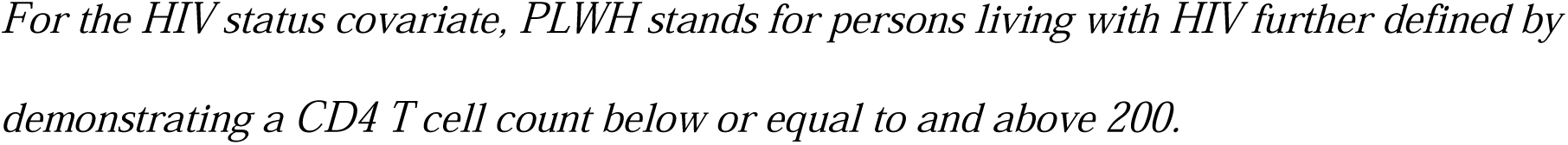
Key characteristics of individuals, households, and influenza viral infections.

|  |  |  |  |  |  |
| --- | --- | --- | --- | --- | --- |
| Individual<br>Characteristic | Year |  | 2016<br>N= 480 | 2017<br>N= 523 | 2018<br>N= 515 |
|  | Site | Agincourt<br>(Rural) | 234 (49) | 261 (50) | 253 (49) |
|  |  | Klerksdorp<br>(Urban) | 246 (51) | 262 (50) | 262 (51) |
|  | Age<br>Group<br>(years) | <5 | 59 (12) | 58 (11) | 59 (11) |
|  |  | 5-11 | 112 (23) | 119 (23) | 139 (27) |
|  |  | 12-18 | 103 (22) | 110 (21) | 85 (17) |
|  |  | 19-40 | 115 (24) | 102 (19) | 142 (28) |
|  |  | >40 | 91 (19) | 134 (26) | 85 (17) |
|  | Sex | Female | 286 (60) | 309 (59) | 325 (63) |
|  |  | Male | 194 (40) | 214 (41) | 190 (37) |
|  | HIV<br>Status | Negative | 403 (84) | 440 (84) | 400 (78) |
|  |  | PLWH CD4<br>≥200 | 64 (13) | 58 (11) | 72 (14) |
|  |  | PLWH CD4<br><200 | 13 (3) | 10 (2) | 22 (4) |
|  |  | Unknown | 0 | 15 (3) | 21 (4) |
| Household<br>Characteristic | Year |  | 2016<br>N = 100 | 2017<br>N = 108 (33) | 2018<br>N = 117 |
|  | Household<br>Size | 3-5<br>Members | 51 (51) | 61 (56) | 82 (70) |
|  |  | 6-10<br>Members | 44 (44) | 41 (38) | 34 (29) |
|  |  | >10<br>Members | 5 (5) | 6 (6) | 1 (1) |
| Observed<br>Infection<br>Episodes | Year |  | 2016<br>N = 174 | 2017<br>N = 239 | 2018<br>N = 210 |
|  | A(H1N1)pdm09 |  | 52 (30) | 4 (2) | 77 (37) |
|  | A(H3N2) |  | 28 (16) | 164 (69) | 3 (1) |
|  | B/Victoria |  | 90 (52) | 0 | 130 (62) |
|  | B/Yamagata |  | 4 (2) | 71 (30) | 0 |

Key characteristics of individuals and households included in the analysis, and influenza virus infections for which shedding kinetics could be estimated. Brackets indicate percentage relative to the given N for the year and category (Individual Characteristic/ Household Characteristic/ Observed Infection Episodes). See Cohen et al. [7] for all observed individuals and infections.
*For the HIV status covariate, PLWH stands for persons living with HIV further defined by* *demonstrating a CD4 T cell count below or equal to and above 200.*

The highest pre-season HAI titers were typically found in individuals between the ages of 10 and 20 years for all the four observed subtypes/lineages (Fig 1). The steepest age gradient in the titers was found between the ages of 0 and 10 years for both influenza A subtypes, while the increase was less pronounced for B/Victoria and spans a broader age range of 0 to 25 years for B/Yamagata. After a peak in individuals in early teenage or adulthood, the titers for all subtypes demonstrated a decline across ages followed by a plateau at older ages around a titer measure of 20-30. In contrast, titer variation started at a high level in early childhood and increased slightly—except for B/Victoria, which declined monotonically, before gradually decreasing with age. This pattern reflects the stochastic nature of infection acquisition and immune boosting in early life, which becomes averaged out as individuals accumulate more exposures over time, as well as intrinsic individual heterogeneity in antibody responses to influenza viruses.

**Fig. 1.**
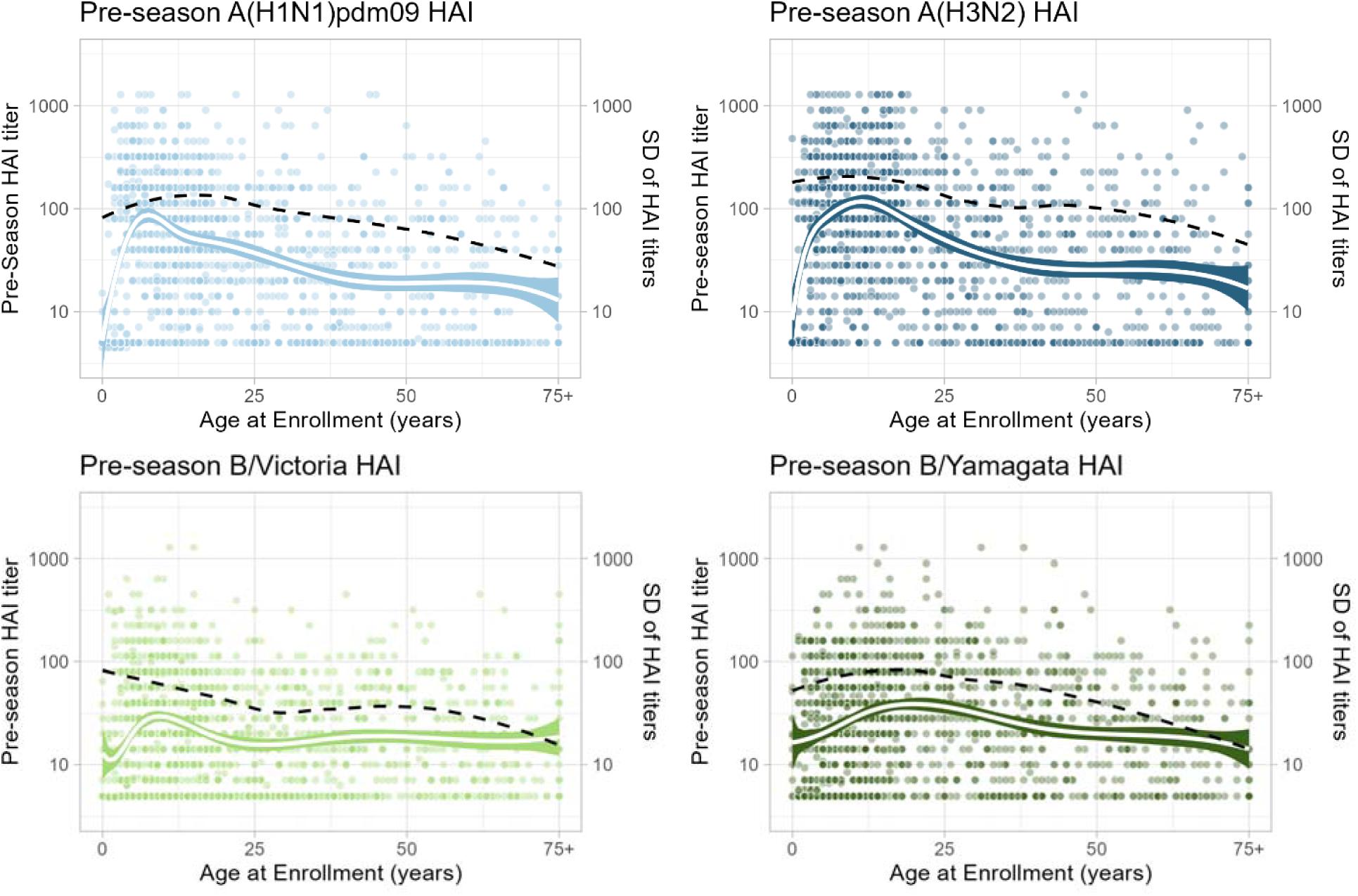
Distribution of the pre-season serum influenza antibody titers. Titers shown as a function of age of the participant at enrollment, separated by subtype/lineage. All participants over 75 are grouped together. The titers were generated using hemagglutination inhibition assay (HAI) against representative circulating viruses during the study period. The white solid line is a fitted linear spline fit with B-spline knots at ages 5, 12, 18, and 40 to describe age-dependent patterns with colored shaded regions representing the 95% confidence intervals. The black dotted line reflects the standard deviation of the titers across ages.

Viral RNA shedding trajectories were estimated for 623 infection episodes, of which 133 (21%) were A(H1N1)pdm09, 195 (31%) were A(H3N2), 220 (35%) were B/Victoria, and 75 (12%) were B/Yamagata (Table 1). Fig 2 summarizes the viral shedding kinetics estimates for each subtype/lineage. The median proliferation duration was similar across subtype (Fig 2B; A(H1N1)pdm09 2.20 days, IQR: 1.98-2.60; A(H3N2) 1.87 days, IQR: 1.62-2.28; B/Victoria 1.78 days, IQR: 1.56-2.16; B/Yamagata 1.86 days, IQR: 1.75-2.26). The median clearance duration was consistently longer than the proliferation duration for all subtypes, and slightly, though not significantly longer, for both type B lineages compared to type A subtypes (Fig 2C; A(H1N1)pdm09 2.52 days, IQR: 1.95-4.47; A(H3N2) 2.57 days, IQR: 2.07-4.92; B/Victoria 3.04 days, IQR: 2.30-5.86; B/Yamagata 3.26 days, IQR: 2.51-5.97). Accordingly, the total durations were similar across subtypes/lineages (Fig 2D; A(H1N1)pdm09 4.7 days, IQR: 4.0-75; A(H3N2) 4.6 days, IQR: 3.8-7.7; B/Victoria 5.2 days, IQR: 4.0-8.4; B/Yamagata 5.3 days, IQR: 4.4-8.4). The coefficients of variation were higher for the clearance duration than proliferation duration, significantly so for all but A(H3N2) by asymptomatic testing for equality of coefficients of variation (p-value < 0.05) ^26^. (Supplementary Table 1). The median estimated peak viral load as proxied by minimum Ct for the episode ranged from 19.8 Ct for A(H3N2) to 22.0 Ct for A(H1N1)pdm09 (Fig 2E; A(H1N1)pdm09 22.0 Ct, IQR: 19.8-23.5; A(H3N2) 19.8 Ct, IQR: 18.4-20.2; B/Victoria 20.5 Ct, 19.3-21.3; B/Yamagata 19.9 Ct, IQR: 17.1-21.1).

**Fig. 2.**
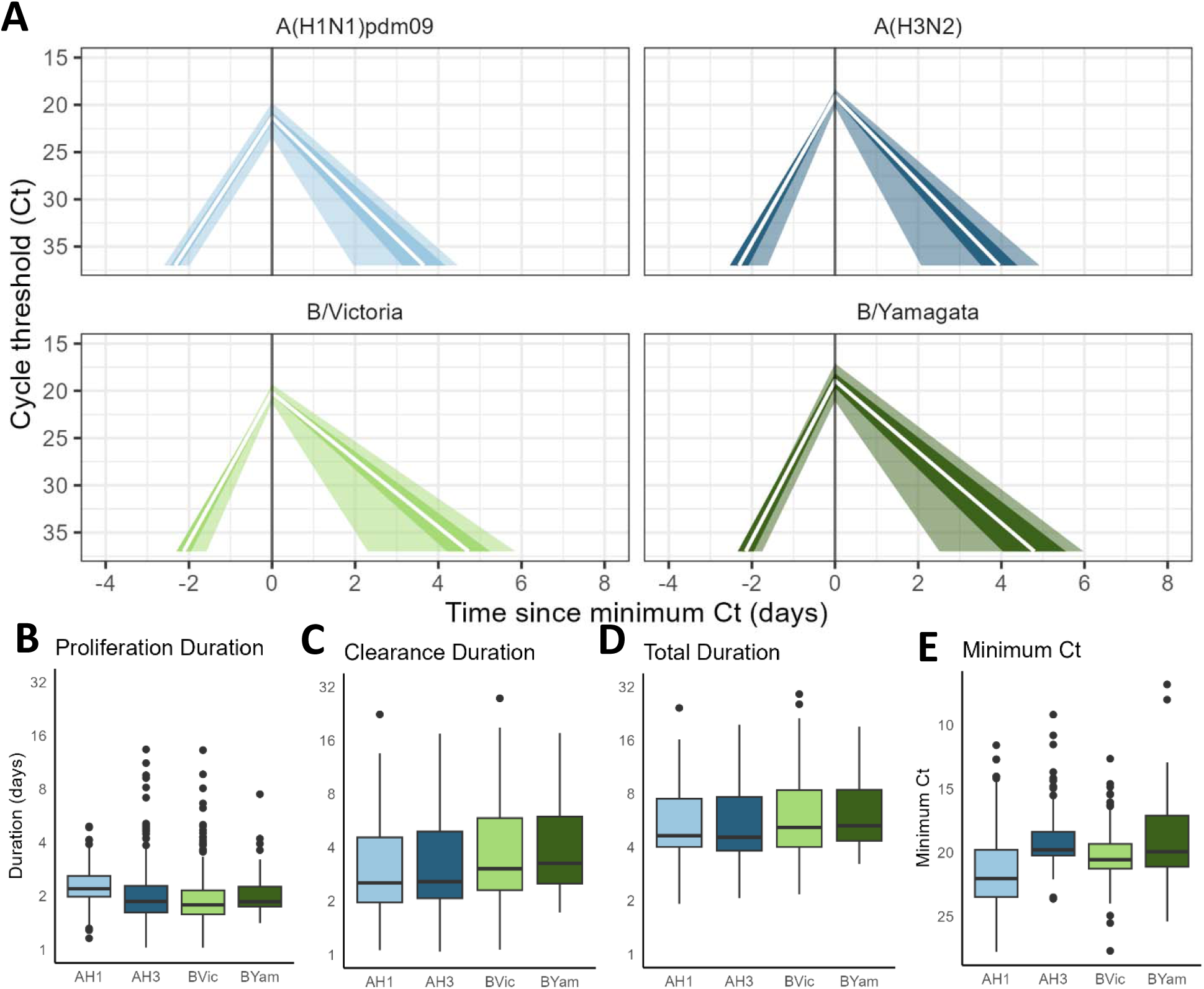
Influenza viral shedding kinetics estimates for each subtype/lineage. (A) Average estimated viral shedding kinetics in Ct values over time since peak shedding (i.e., time since the minimum Ct value). The white solid line represents the posterior mean estimate from the Bayesian piecewise linear viral shedding model, the darker shaded area represents the 95% confidence intervals, and the lighter shaded area represents the interquartile range. (B) Distributions of the proliferation duration estimates for each subtype/lineage. (C-E) same as (B) for clearance duration, total duration, and minimum Ct (peak viral load), respectively.

### Age and pre-season HAI titers association with the shedding kinetics of seasonal influenza

Fig 3 shows predictors of the total duration and the peak viral load (minimum Ct) of viral RNA shedding by influenza subtype/lineage. For A(H1N1)pdm09 infections, higher pre-season HAI titers correlated with shorter durations of shedding, with a 2.3-day reduction in duration time for those with a titer of at least 40 compared to those with lower titers [95% confidence interval (CI): −3.90 - −0.41 days]. This was not seen for other subtypes/lineages. Younger individuals tended to shed for longer, even after adjusting for HAI titers. In particular, children <5 years old exhibited a 3-5 day longer shedding duration for all subtypes/lineages apart from B/Yamagata compared to persons >40 years old (A(H1N1)pdm09 3.33 days, 95% CI: 0.87-5.79; A(H3N2) 4.16 days, 95% CI: 1.72-6.61; B/Victoria 4.46 days, 95% CI: 1.24-7.69). Children aged 5-11 years also demonstrated longer durations for infections of A(H1N1)pdm09 [2.67 days, 95% CI: 0.21-5.13] and A(H3N2) [2.64 days, 95% CI: 0.65-4.64]. HIV status, even among the immunocompromised, was not significantly associated with the total duration of shedding for any of the influenza subtypes. Regarding peak viral load, no significant association was found between any of the included covariates and viral load reduction for any subtype or lineage (Fig 3, bottom panel).

**Fig. 3.**
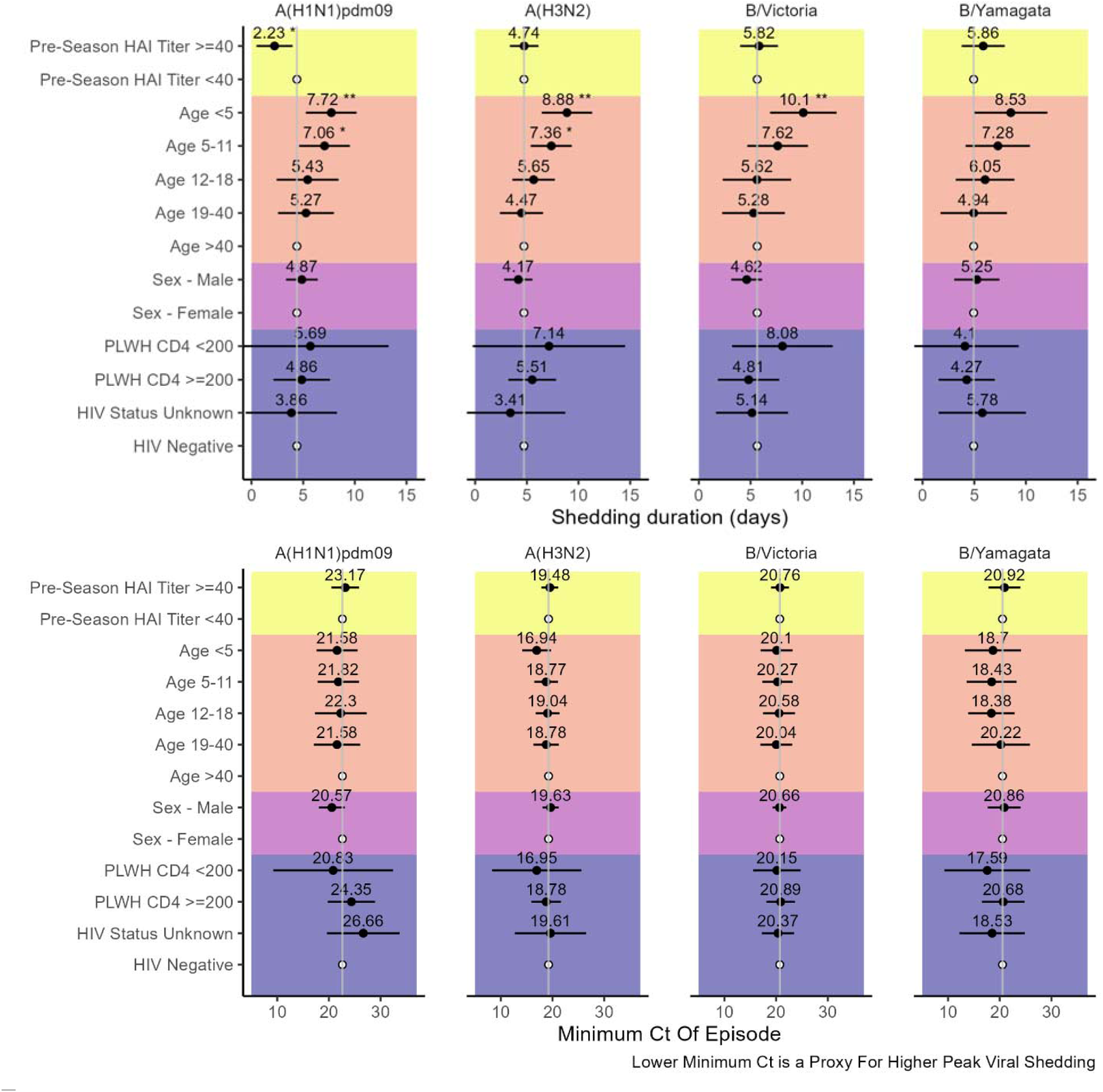
Predictors of influenza virus shedding characteristics by subtype/lineage based on multivariable linear regression. The top panel regresses the total shedding duration and the bottom panel the peak viral load, proxied by the minimum Ct of the infection episode. For the HIV status covariate, PLWH stands for persons living with HIV further defined by demonstrating a CD4 T cell count below or equal to and above 200. For shedding duration, estimates are given in units of days with lines representing the 95% CI. The central line indicates the mean duration for a given subtype/lineage. The reference groups are designated by a white dot. For the shedding duration the reference estimates for each subtype are as follows: A(H1N1)pdm09 4.39 days, A(H3N2) 4.71 days, B/Victoria 5.64 days, B/Yamagata 4.94 days. For the minimum Ct of episode, the reference estimates are A(H1N1)pdm09 22.62 Ct, A(H3N2) 21.06 Ct, B/Victoria 20.54 Ct, B/Yamagata 20.72 Ct. Asterisks denote p-value: ‘***’: <0.001 ‘**’: <0.01 ‘*’: <0.05 calculated via Satterthwaite’s degrees of freedom method.

### Household and community influenza exposures, pre-season HAI titers, and age jointly shape the risk of influenza acquisition

Next, we used the shedding data to inform a daily transmission model accounting for time-varying influenza exposure in the household. We also considered time-varying infection risk from the community, and demographic and immunological characteristics of household members, using a multivariable logistic regression framework (see Fig 4 and methods for details on the approach, and Fig 5 for results). We find a strong effect of the household force of infection (FOI, proxied by viral shedding) on risk of infection acquisition, where a 10 Ct unit increase in viral load within the household associated with a 65-95% increase in risk depending on subtype/lineage (A(H1N1)pdm09 hazard ratio (HR), 1.96 95% CI: 1.58-2.44; A(H3N2) 1.64 HR, 95% CI: 1.32-2.03; B/Victoria 1.77 HR, 95% CI: 1.42-2.20; B/Yamagata 1.70 HR, 95% CI: 1.15-2.52). The community FOI (proxied by influenza infection prevalence in the cohort) was significantly associated with an increase in risk of infection acquisition for all four subtypes/lineages, such that a 0.01 increase in community influenza prevalence in the population, for which the peak prevalence was between 5-6%, was associated with a 70-100% increase in risk (A(H1N1) pdm09 1.89 HR, 95% CI: 1.68-2.13; A(H3N2) 1.69 HR, 95% CI: 1.56-1.82; B/Victoria 2.01 HR, 95% CI: 1.83-2.20; B/Yamagata 1.95 HR, 95% CI: 1.74-2.19; supplemental figure 10).

**Fig. 4.**
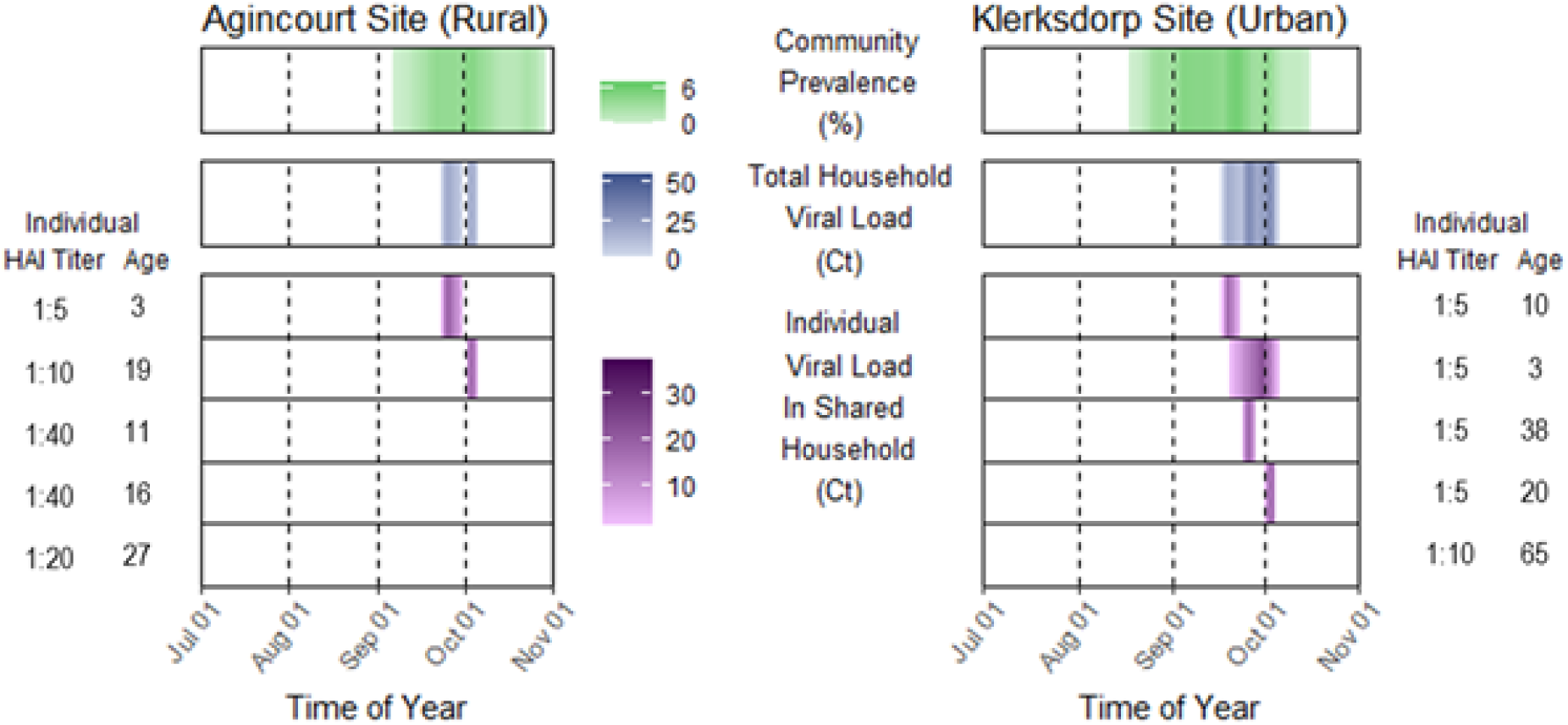
Visualization of key variables of the influenza household transmission model. Depicts the interplay between community prevalence, household exposure, pre-season titers and age. Two individual households, selected from rural (left) and urban (right) communities, are presented. Influenza community prevalence (top panel) is shown as a percentage of the enrolled cohort with an observed PCR-confirmed infection. The total influenza household exposure (second panel) is shown as a composite of individual viral loads Ct values (bottom panel) within the household. The ages (in years) and HAI titers of household members are indicated on either side. The model is run on a daily time scale.

**Fig. 5.**
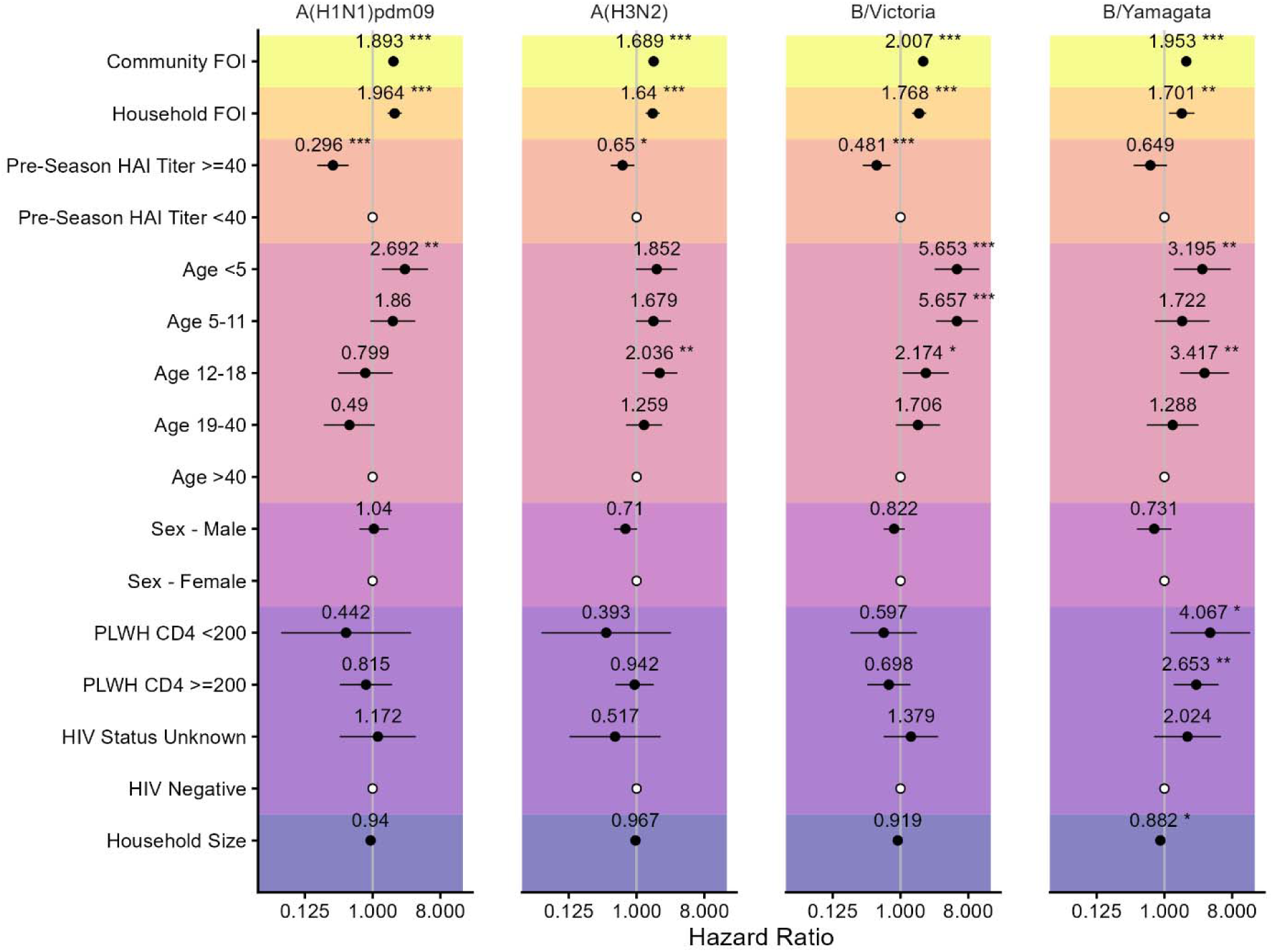
Factors influencing the risk of influenza virus infection acquisition by subtype/lineage based on multivariable logistic regression. Variables include the community force of infection (FOI), household FOI, pre-season HAI titer, age, sex, HIV status, and household size. Community FOI is indicated with respect to a 0.01 increase in absolute community prevalence (defined as the total number of infection episodes within the cohort at a given time / cohort population). Household FOI is indicated with respect to a step by 10 Ct units within the household. For the HIV status covariate, PLWH stands for persons living with HIV further defined by demonstrating a CD4 T cell count below or equal and above 200. Black dots represent point estimates of hazard ratios (logistic regression is equivalent to a Cox model in this context), with back horizontal lines indicating 95% CI intervals. The reference groups for categorical variables are designated by a white dot. Asterisks denote p-value ‘***’: <0.001 ‘**’: <0.01 ‘*’: <0.05.

High pre-season HAI titers were protective against infection for all subtypes with the exception of B/Yamagata, with a titer of at least 1:40 providing a 50-70% reduction in risk of infection acquisition (Fig 5; A(H1N1) pdm09 0.30 HR, 95% CI: 0.18-0.48; A(H3N2) 0.65 HR, 95% CI: 0.45-0.93; B/Victoria 0.48 HR, 95% CI: 0.31-0.74). Even after controlling for the household and community FOI as well as pre-season HAI titers, there was still a residual association between age and risk of infection acquisition. Compared to the reference group of ≥40 years old, children had an increased risk of infection, although the specific age subgroup exhibiting higher susceptibility depended on the subtype. Those <5 years old had an increased risk of infection by 170% to 460% for all subtypes except A(H3N2) (A(H1N1)pdm09 2.69 HR, 95% CI: 1.33-5.46; B/Victoria 5.65 HR, 95% CI: 2.85-11.21; B/Yamagata 3.19 HR, 95% CI: 1.33-7.65). Those aged 5-11 years demonstrated a significantly increased risk of infection acquisition compared to the reference group for B/Victoria (B/Victoria 5.66 HR, 95% CI: 2.98-10.73); whereas those in the 12-18 year old group had a higher risk for all subtypes but A(H1N1)pdm09 (A(H3N2) 2.04 HR, 95% CI: 1.19-3.49; B/Victoria 2.17 HR, 95% CI 1.07-4.42; B/Yamagata 3.42 HR, 95% CI 1.61-7.23).

For B/Yamagata infections, larger household size was associated with a decreased risk of infection acquisition, such that one additional person in the household contributed about 10% reduction in risk (0.88 HR, 95% CI 0.80-0.98). For B/Yamagata, compared to individuals that were HIV negative, people living with HIV (PLWH) displayed an increased risk of infection acquisition that scaled with the level of immuno-suppression (PLWH CD4<200 4.07 HR, 95% CI: 1.20-13.86; PLWH CD4>=200 2.65 HR, 95% CI: 1.33-5.28). No other measured characteristic was associated with a significant change in risk of infection acquisition (p-value > 0.05).

## Discussion

We analyzed data from the PHIRST cohort in South Africa to refine our understanding of the viral, demographic, and immunologic factors that influence influenza virus transmission. The PHIRST study had a unique design with high frequency real-time reverse transcription polymerase chain reaction (rRT-PCR) sampling at intervals similar to the timescale of influence virus transmission, enabling us to characterize the shedding kinetics of each observed infection episode. In this cohort that comprised a young population with a relatively high prevalence of comorbidities, we demonstrate that age is a stronger predictor of shedding duration than pre-season HAI titers. From the estimated shedding kinetics of household members and viral activity in the cohort, we model the temporal dynamics of influenza exposure within the household and community, both of which are predictive of risk of infection acquisition. Further, we find that pre-season HAI titers are a correlate of protection for the predominant influenza subtypes/lineages (A/H3N2, A/H1N1, B/Victoria); however, even after adjusting for titers, children and adolescents retained higher susceptibility to influenza for all subtypes/lineages.

Our results indicate that viral shedding intensity among household contacts was predictive of the risk of acquiring influenza infection. Specifically, a 10-unit increase in shedding intensity, as indicated by the Ct value, was associated with approximately a 95% increased risk for A(H1N1)pdm09, 65% for A(H3N2), 80% for B/Victoria, and 70% for B/Yamagata. This observed difference between subtypes and lineages may reflect variation in the infectivity of viral particles, where the same viral load (Ct) corresponds to differing numbers of infectious virions due to subtype-specific differences in replication efficiency, particle stability, and host immune responses not fully captured by controlling for HAI titer and age. These findings are consistent with human challenge studies that have demonstrated a correlation between the risk of influenza virus infection acquisition and challenge viral dose ^1,2^. In contrast, previous case-ascertained household transmission studies did not observe significant correlations between the viral shedding intensity of the index case and subsequent infectivity ^5,12,18^. These studies concentrated on post-symptomatic viral shedding and thereby could not evaluate pre-symptomatic and asymptomatic shedding, as well as exposures to multiple infected household members. Additionally, the PHIRST cohort included a substantial proportion of children, whose influenza infection episodes were significantly longer and exhibited higher peak viral loads compared to adults, after controlling for pre-season HAI titer. The presence of these prolonged shedding episodes provided the opportunity to examine infectivity hazards with a greater temporal variability than prior studies, thereby enhancing the statistical power of our analysis. Similarly, increased prevalence in the general community, estimated from the overall cohort population, accounting for any exposure outside of the household such as in schools or workplaces, was also strongly correlated with an increased risk of infection acquisition.

Controlling for time-varying influenza exposures within the household and from the general community, we found that HAI titers of at least 1:40 were a correlate of protection against influenza virus infection for all strains except for B/Yamagata, conferring at most 70% protection against A(H1N1)pdm09 and at least 35% protection against A(H3N2). This relationship between pre-existing immunity and risk of infection acquisition aligns well with prior studies that have identified HAI titers of at least 1:40 as providing approximately 50% reduction in risk, even though this cutoff was based on adults with vaccine-mediated immunity while the PHIRST cohort was focused on natural immunity ^21,22^. Titers against B/Yamagata did not appear to confer protection when considering a threshold of 1:40. However, running the same transmission model, with the only difference being to treat titers as a continuous variable rather than ≥40 or <40 categorical, higher B/Yamagata titers did associate with a reduce infection risk (Supplementary Fig 8). These results suggest either that the threshold for protection may be higher than 1:40 for this lineage, or that there was a lack of power due to B/Yamagata causing fewer infections than the other subtypes/lineages in our dataset. Interestingly, the effect size of A(H3N2) is much smaller than that of A(H1N1)pdm09 and when evaluating the titer on a continuous basis on a log base four scale, the titer is no longer significantly associated with a reduction in risk. Putative explanations include antigenic variation in the circulating strain of A(H3N2) that did not match pre-existing antibody titers, or immune distinctiveness of A(H3N2). As it relates to age, the observed titer patterns for influenza A indicate a rapid establishment of strain-specific antibody responses during early childhood, consistent with strong early-life imprinting to influenza A antigens ^27^. In contrast, the broader and less pronounced gradient observed for influenza B likely reflects less frequent immune boosting due to lower infection rates, as well as the inferior persistence of antibodies against influenza B viruses compared with influenza A ^28,29^.

A key result of our study was the strong residual effect of age on the risk of infection acquisition, after adjustment for HAI titers. Previous research has indicated that children have a higher risk of infection for both type A and B influenza ^14,30^. Compared to older individuals, children are thought to experience higher contact rates, more exposure-prone behavior, and relative immunological naivety beyond that measured by HAI titers. Our data can shed light on the putative contribution of each of these hypotheses. If contact rates and other behavioral risks were the dominant factors in the age variation in infection risk, there should be consistent age relationships across all subtypes, as contact rates and behavior universally affect infection transmission. Kleynhans et al. ^31^ conducted a cross-sectional study within the PHIRST cohort and found that children aged 14-18 years had the highest contact rates of all participants. This could partially explain the heightened risk of infection acquisition observed in individuals aged 12-18 years for A(H3N2), B/Victoria, and B/Yamagata, but it does not support the increased risk seen in those under 11 years old for A(H1N1)pdm09, B/Victoria, and B/Yamagata. While there could be other age-specific factors such as contact rates and behavioral heterogeneities unaccounted for, our data suggests a potential residual effect of age-specific immune responses. Previous work has suggested that the 1:40 HAI titer cutoff is inaccurate to use as a correlate of protection when evaluating vaccines for children ^23^. While our household model accounts for variations in pre-existing HAI titers both in categorical and continuous scales, other age-dependent immune mediators that are not captured by HAI titers likely play a role (Supplementary Figs 7 and 8). Since children are considered high-transmitters and a key group for vaccination, it will be important to identify correlates of protection against influenza infection that are age-appropriate ^30,32^.

We found a significant gradient of influenza virus shedding duration with age across all subtypes/lineages in our adjusted models, in which the youngest children shed the longest. We did not find significant impact of pre-season HAI titer level on shedding duration, except for influenza A(H1N1)pdm09. Since children, nevertheless, shed the longest, it is possible that unobserved pre-existing components of adaptive immune memory, such as non-HAI antibodies, could have reduced the shedding duration. These additional immune functions likely operate on viral clearance, as the coefficient of variation was significantly higher for clearance duration than proliferation duration for all but A(H3N2) (Supplementary Table 1). If immune functions beyond HAI antibodies, such as those targeting neuraminidase, the HA stalk, or other influenza-specific cellular responses, play a more critical role in promoting viral clearance for A(H3N2), B/Victoria, and B/Yamagata infections, while pre-existing HAI antibodies are more effective at preventing infection and correlate more closely with viral control for A(H1N1)pdm09, this could explain the subtype-specific effects observed on shedding duration ^33^. This interpretation aligns with the hypothesis that pre-season HAI antibodies primarily act to block infection rather than to reduce viral shedding once infection is established, with the exception of A(H1N1)pdm09, where HAI titers may better reflect protective immune responses involved in limiting viral replication.

Further, this hypothesis could in part explain why there was no significant association between titers above 40 and viral load reduction. But it should be noted that there was also no residual effect of age, and it is also likely that since peak viral load is less directly measurable, it is likely less accurately estimated and further interpretation is limited. More broadly, the endemicity of seasonal influenza implies the number of influenza exposures accumulates with age, and so would immune memory. Overall, immune compartments other than serum HA antibodies, such as neuraminidase or stalk antibodies, influenza-specific B-cell and T-cell memory, or binding antibodies that do not inhibit hemagglutination, may contribute to the prevention of infection acquisition and/or the reduction of viral shedding ^2,33–35^.

We also explored other individual and household-level characteristics associated with both the duration of viral shedding and the risk of infection acquisition. Previous studies have indicated that HIV status influences the risk of morbidity and mortality from influenza-related illness ^36,37^. While there was a relatively large portion of people living with HIV included in our analysis, most participants living with HIV were on effective treatment, limiting the power of analysis on immunocompromised individuals. We found no significant correlation between HIV status, including PLWH who were immunocompromised, and the duration of viral shedding. However, participants living with HIV had an increased risk of infection acquisition with the B/Yamagata lineage virus. Additionally, for B/Yamagata, individuals living in larger households exhibited a lower risk of infection after controlling for household FOI. This finding is consistent with previous influenza studies that have suggested a “dilution effect” of household contact, wherein members of larger households experience a reduced per-contact transmission risk compared with those in smaller households ^38^. However, this does not necessarily imply a lower overall household infection risk, as we observed that the cumulative household infection risk (defined as the fraction of household members infected during a given season) varied across households of different sizes (Supplementary Fig 11).”

Our analysis has several limitations. First, despite the extensive rRT-PCR testing scheme, there were still 2–3-day intervals between sample collections. This could result in missing very short infection episodes, potentially leading to missed influenza infections. We also did not use quantitative PCRs but rather used the Ct value as an indirect measure of viral load. Second, our analysis assumes that pre-season HAI titer measurements remain constant throughout each influenza season, despite the knowledge that HAI titer might wane over the season. Third, we were unable to evaluate the impact of co-circulating influenza viruses as we excluded instances of coinfection from our analysis. Fourth, our transmission model lacks a specific contact structure, meaning it does not account for heterogeneity in interactions among household members or demographic differences in community contact exposure ^39^. As a result, we cannot fully resolve the contribution of behavioral and biological risk factors to age differences in susceptibility, although subtype differences point at possible immunological factors (related to the amount of antigenic drift and number of past exposures associated with each lineage). Furthermore, the absence of influenza vaccination in our cohort limits the applicability of our findings to settings with greater vaccine usage, although it allows us to focus on the role of natural immunity. Additionally, the number of observed B/Yamagata infections was lower compared to other subtypes limiting the statistical power for this lineage. It is worth noting that globally, the B/Yamagata lineage is now considered extinct in the post-COVID-19 pandemic period ^40^. Finally, our study was limited to the pre-COVID-19 period; it remains unclear how the immunity gap produced by the lack of influenza circulation during the COVID-19 pandemic would impact the relationship between pre-existing HAI titers, age, and the risk of infection ^10,41,42^.

In summary, by modeling high-resolution virological samples from a detailed cohort study, we were able to reconstruct the time-varying intensities of influenza exposures within households and from the general community, both of which significantly correlated with the risk of influenza infection acquisition. After adjusting for these exposures, we found that pre-season HA antibodies reduced the risk of infection acquisition by influenza A(H1N1)pdm09, A(H3N2), and B/Victoria, but not B/Yamagata and reduced shedding duration for A(H1N1)pdm09. After adjustment for HAI titers, we found a residual effect of age on shedding duration and infection risk. Furthermore, the residual age effects on susceptibility and shedding duration and the inconsistency of the observed age trend between subtypes highlight the need for more work on the roles of immune compartments beyond serum HA antibodies, such as antibodies to neuraminidase and HA stalk, B-cell, T-cell, neutralization and other effector functions of antibodies as potential correlates of protection against influenza.

## Data Availability

The study protocol including informed consent forms is available on the NICD website (https://www.nicd.ac.za/wp-content/uploads/2021/02/PHIRST-SARS-CoV-2-protocol-V1-amendment-Nov2020-incl-upd-consent.pdf). The investigators welcome enquiries about possible collaborations and requests for access to the data set. Data will be shared after approval of a proposal and with a signed data access agreement. Investigators interested in more details about this study, or in accessing these resources, should contact the principle investigator, Prof Cheryl Cohen, at NICD.

## Acknowledgements

The authors would like to thank all the individuals who kindly agreed to participate in the study as well as the PHIRST group. Data collection of the PHIRST flu study was supported by the National Institute for Communicable Diseases of the National Health Laboratory Service (https://www.nicd.ac.za/) and the US Centers for Disease Control and Prevention (https://www.cdc.gov/) (cooperative agreement number 5U51IP000155). MS would like to acknowledge support from Princeton University’s Office of Undergraduate Research Undergraduate Fund for Academic Conferences through the President’s Fund. BG would like to acknowledge support from Princeton Catalysis and Princeton Precision Health. The findings and conclusions in this report are those of the authors and do not necessarily represent the official position of the funding agencies, the NIH or the U.S. Centers for Disease Control and Prevention.

## Author contributions

Conception and design of the study: MS, ST, CV, KS, CC

Data collection and laboratory processing: JK, JM, FkT, OH, AvG, NW, AB, LM, NaM, KK, LL, KM, FW, FxG, ST, CC

Analysis and interpretation: MS, JK, JM, MlM, AvG, NW, ST, BG, CV, KS, CC

Drafting or critical review of the article: MS, JK, JM, MlM, FkT, OH, AvG, NW, AB, LM, NaM, KK, LL, KM, FW, FxG, ST, BG, CV, KS, CC

## Inclusion & Ethics Statement

The PHIRST protocol was approved by the University of the Witwatersrand Human Research Ethics Committee (Reference 150808) and the US CDC’s Institutional Review Board relied on the local review (#6840). The protocol was registered on http://clinicaltrials.gov on 6 August 2015 (Reference NCT02519803). Participants provided individual written consent or assent prior to enrollment and received a grocery store voucher of ZAR25-30 (USD 2–2.5) per visit for their time. (See 45 C.F.R. part 46.114; 21 C.F.R. part 56.114).

## Data Availability

To access data, including individual participant data and a data dictionary defining each field in the dataset, please submit a proposal to C.C. These data can be made available through a data access agreement or material transfer agreement.

## Code Availability

All the statistical analysis for this study and the figures were performed in R (version 4.3.1, RStudio 2023.6.2.561) and the corresponding code with mock data can be found at https://github.com/msauter19/phirst-flu-shed-trans.

## Supplementary Information

Supplementary text, tables, and figures

