## Supplementary text, tables, and figures for "The roles of pre-season immunity, age, viral shedding, and community exposures in shaping influenza household transmission dynamics"

### Supplementary Methods

#### 1. Spline-Based Visualization of Age-Dependent Trends

The age-dependent trend line of HAI titers in Fig 2 and Supplementary Fig 9 were generated in R (version 4.3.1, RStudio 2023.6.2.561) using the base R package splines to create spline-based fits to capture non-linear trends in the data. These curves were constructed with B-splines, with knot placement at ages 5, 12, 18, and 40 to describe age-dependent patterns. We used a cubic spline (degree = 3) because cubic functions are the standard choice in spline modeling; they ensure smooth transitions (continuous first and second derivatives) between segments while maintaining interpretability.

#### 2. Characterizing kinetics of influenza viral shedding

##### 2.1 Defining subtype-specific infection episodes

An infection episode was defined as a series of consecutive positive samples from the same individual for a given influenza subtype or lineage, as previously described by Cohen et al.[1]. Briefly, following the first positive sample, an episode included all subsequent samples until either (i) more than two weeks elapsed between two consecutive positive samples, or (ii) a sample tested positive for a different subtype than the preceding one. Negative samples occurring within an episode were retained, as transient negative results can arise from fluctuations in viral load or variable sample quality.

In some cases, samples tested negative for all subtype-specific primers despite being positive for the broader influenza type A or B primers. When such samples occurred within the timeframe of an established infection episode for a known subtype, they were retained and assigned the same subtype as the surrounding samples. Samples that remained untyped after this correction were assigned a subtype based on the dominant circulating lineage during that season: B/Victoria in 2016, A(H3N2) and B/Yamagata in 2017, and B/Victoria and A(H1N1)pdm09 in 2018. Non-subtyped samples likely reflected low viral RNA concentrations and excluding them could bias population-level shedding parameter estimates. However, due to the co-circulation of A(H3N2) and A(H1N1)pdm09 in 2016, non-subtyped influenza A samples from that year were excluded. This exclusion affected 14 samples, each representing a distinct infection episode.

To characterize the viral shedding kinetics, we modeled the Ct values from the raw serial RT-PCR data for each observed episode. We obtained up to three different Ct values for one sample, one for the detection assay and one for each subtype that was detected in the case of coinfections. Though the Ct values of the assays typically correlate with each other, the different assays had different calibrations and cannot be directly compared. Generally, it is assumed that the subtype assay is more sensitive to the true RNA concentration in the sample, as it more precisely identifies the present RNA. However, given our interest of including positive non-subtyped samples for the analysis (those samples that tested negative and were typed by the detection assay, but had a Ct value over the threshold on the subtype assays) all the raw Ct data observations were taken from the FTD Flu/RSV detection assay (Fast Track Diagnostics, Luxembourg) [2,3].

#### 2.2 Modeling Viral Shedding Kinetics Using a Hierarchical Bayesian Framework

As previously described by Kissler et al [4,5], this viral shedding model assumes that the proliferation phase, from the onset of infection to the peak viral load, can be represented by an exponential increase in viral RNA concentration, and that the subsequent clearance phase, from the peak to the end of infection, follows an exponential decline. Consequently, cycle threshold (Ct) values measured by RT-PCR, which serve as a proxy for the negative logarithm of viral RNA concentration, are expected to decrease linearly from the limit-of-detection threshold ($Ct_{lod}$=37) to a minimum Ct value ($Ct_{min}$, representing peak viral RNA concentration) during the proliferation phase, and then increase linearly back to $Ct_{lod}$ during the clearance phase. For convenience, we define

$$\Delta Ct(t) = Ct(t) - Ct_{lod}$$

which represents viral RNA shedding intensity over time, proportional to the logarithm of viral RNA load. The peak shedding intensity is therefore given by

$$\Delta Ct_{max} = Ct_{lod} - Ct_{min}$$

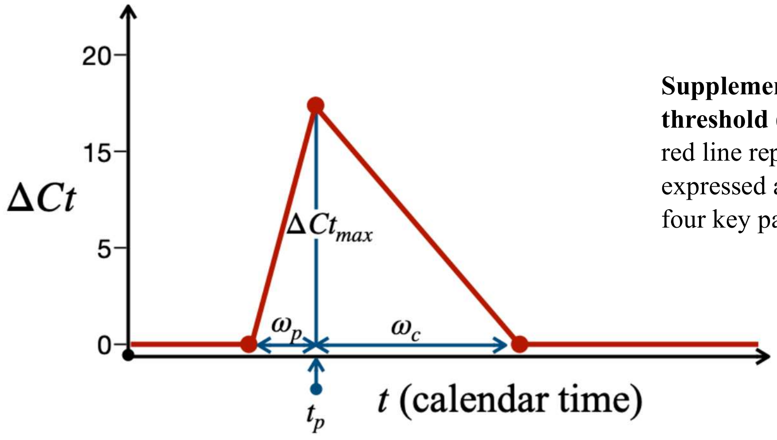


**Supplementary Fig. 1. Theoretical trajectory of cycle threshold (Ct) values during an infection episode.** The red line represents the modeled viral shedding trajectory, expressed as $\Delta Ct(t)$ over time. The blue lines indicate the four key parameters estimated in the model:

Under these assumptions, individual viral shedding kinetics can be described as a linear increase in ΔCt from 0 to the peak shedding intensity ($\Delta Ct_{max}$) during the proliferation phase, followed by a linear decrease from $\Delta Ct_{max}$ back to 0 during the clearance phase. Supplementary Fig. 1 illustrates this theoretical kinetic profile, representing the model expectation E[ΔCt(t)] , and visualizes the key parameters estimated in the shedding model: $\omega_p$ for the proliferation duration, $\omega_p$ for the clearance duration, $t_p$ for the calendar timing of peak viral shedding, and $\Delta Ct_{max}$ for the peak shedding intensity. Under this viral shedding framework, the expected individual shedding trajectory E[ΔCt(t)] can thus be expressed as a set of piecewise linear equations.

$$E[\Delta Ct(t)] = \begin{cases} 0, & t \leq t_p - \omega_p \\ \dfrac{\Delta Ct_{max}}{\omega_p}\left(t + \omega_p - t_p\right), & t_p - \omega_p < t \leq t_p \\ \Delta Ct_{max} - \dfrac{\Delta Ct_{max}}{\omega_c}, & t_p - \omega_p < t \leq t_p \\ 0, & t > t_p + \omega_c \end{cases}$$

We then specified an observation model, assuming that the observed ΔCt(t) values are drawn from a zero-truncated normal distribution with mean E[ΔCt(t)] and standard deviation $\sigma$, where $\sigma$ represents the observational noise shared across all infections episodes of a given subtype.

$$\Delta Ct(t) \sim \max\left(0, \mathrm{Normal}(E[\Delta Ct(t)], \sigma)\right)$$

For each of the individual parameters ($\omega_p$, $\omega_c$, and $\Delta Ct_{max}$ ), excluding $t_p$ which does not represent intrinsic biological process of viral replication, we implemented a Bayesian hierarchical model at the population level to constrain individual estimates within biologically plausible ranges. Specifically, each individual parameter was assumed to be drawn from a population-level lognormal distribution, defined as follows:

$$\ln(\omega_p) \sim \text{Normal}\left(\ln\left(\mu_{\omega_p}\right), \ln\left(\sigma_{\omega_p}\right)\right)$$

$$\ln(\omega_c) \sim \text{Normal}\left(\ln\left(\mu_{\omega_c}\right), \ln\left(\sigma_{\omega_c}\right)\right)$$

$$\ln(\Delta Ct_{max}) \sim \text{Normal}\left(\ln\left(\mu_{\Delta Ct_{max}}\right), \ln\left(\sigma_{\Delta Ct_{max}}\right)\right)$$

where $\ln(\cdot)$ denotes the natural logarithm function. To improve computational efficiency, constrained the parameters to biologically plausible upper bounds: $\omega_p < 15\, days$, $\omega_c < 30\, days$, and $\Delta Ct_{max} < 37$, as values beyond these thresholds are considered implausible for influenza viral kinetics.

The priors of each of the population level mean parameters ($\mu_{\omega_p}$, $\mu_{\omega_c}$, $\mu_{\Delta Ct_{max}}$) were specified as truncated normal distributions covering biologically plausible ranges of variation in viral proliferation duration ($\omega_p$), clearance duration ($\omega_c$), and peak viral shedding ($\Delta Ct_{max}$). These priors, were informed primarily by published viral kinetics models and relevant influenza human challenge studies [1,6,7], specifically:

$$\text{Prior}\left(\mu_{\omega_p}\right) \sim \text{Normal}(2, 1), \mu_{\omega_p} > 0$$

$$\text{Prior}\left(\mu_{\omega_c}\right) \sim \begin{cases} \text{Normal}(4, 3) \text{ for influenza type A}, \mu_{\omega_c} > 0 \\ \text{Normal}(5, 3) \text{ for influenza type B}, \mu_{\omega_c} > 0 \end{cases}$$

$$\text{Prior}\left(\mu_{\Delta Ct_{max}}\right) \sim \text{Normal}(37/2, 37/6), \mu_{\Delta Ct_{max}} > 0$$

The priors for the corresponding population-level variation parameters ($\sigma_{\omega_p}$, $\sigma_{\omega_c}$, $\sigma_{\Delta Ct_{max}}$) are selected to reflect plausible ranges of biological variability observed across individuals:

$$\text{Prior}\left(\sigma_{\omega_p}\right) \sim \text{Normal}(1, 1/2), \sigma_{\omega_p} > 0$$

$$\text{Prior}\left(\sigma_{\omega_c}\right) \sim \text{Normal}(3, 3/2), \sigma_{\omega_c} > 0$$

$$\text{Prior}\left(\sigma_{\Delta Ct_{max}}\right) \sim \text{Normal}(37/6, 12), \sigma_{\Delta Ct_{max}} > 0$$

The peak timing ($t_p$) of each individual shedding episode was estimated independently at the individual level, as it is primarily influenced by individual's exposure to household and community forces of infection rather than by intrinsic within-host viral replication dynamics. Specifically, the prior distribution for each individual's peak timing was assumed to follow a normal distribution centered on the timing of their observed highest $\Delta Ct$ value, with a standard deviation of 2 days to reflect the empirical sampling interval.

$$\text{Prior}\left(t_p\right) = \text{Normal}\left(t_p^{obs}\left(\Delta Ct\left(t_p^{obs}\right) = \Delta Ct_{max}^{obs}\right), 2\right)$$

This choice reflects the empirical sampling frequency of approximately 2–3 days between consecutive respiratory sample collections, allowing sufficient flexibility for the model to capture inter-individual variation in peak timing while remaining consistent with the temporal resolution of the data.

Given the above model specification, we excluded certain infection episodes from the shedding model estimates due to the proximity of missing samples. This included if there were two missing samples prior to the first positive sample or after the last positive sample of an episode. In other words, that we did not have data for a week on either end of the infection, inhibiting the opportunity to estimate the duration with accuracy. We also excluded infections that were cut off by either the start or the end of the collection period, with a positive occurring within one negative or missing sample from start or end collection date; this was mostly of the B/Victoria wave that occurred relatively late in 2018. This excluded 58 out of 681 infections.

The Bayesian hierarchical models described above were implemented in Stan (rstan version 2.32.6) and interfaced through R (version 4.3.1, RStudio 2023.6.2.561). Separate models were fitted for each influenza subtype or lineage using four Markov Chain Monte Carlo (MCMC) chains, each run for 1,000 iterations. The estimated $\sigma$ for each influenza subtype reflected the typical deviation of observed data from model predictions (A(H1N1)pdm09 median = 0.65, 95% CI: 0.60–0.73; A(H3N2) median = 1.00, 95% CI: 0.96–1.05; B/Victoria median = 1.25, 95% CI: 1.14–1.39; B/Yamagata median = 0.96, 95% CI: 0.89–1.30). Bayesian modeling of influenza viral shedding was performed using Stan, and posterior summaries and model diagnostics were obtained through the rstan package.

For downstream regression analyses (described in the next section), we accounted for uncertainty arising from potential censoring and missing observations by resampling from the posterior distributions. Each regression was repeated 100 times, drawing one random posterior estimate per individual from the shedding model outputs, and the resulting regression estimates were pooled to obtain the final reported values.

#### A note on the shedding model ill-fitted estimates

To evaluate the model fit, we analyzed the number of divergent transitions for each of the four model runs, relevant to the numerical instability in Hamilton Monte Carlo sampling and should ideally be zero, and the Gelman R-hat statistic for each posterior, a convergence diagnostic where values close to 1 indicate good mixing across chains. While the majority of fits demonstrated satisfactory convergence and stability – across all model runs there were no divergent transitions – a subset exhibited mild deviations from ideal diagnostic criteria.

At the population level, 3 (36%) of the 24 posteriors had a Gelman R statistic that exceeded the 1.1 threshold. These were for the proliferation mean posteriors for A(H3N2), and both the proliferation mean and clearance posteriors for B/Victoria, which might have been a result of the relatively large observed variation in duration. At the individual-episode level (n = 623), 15 (2%) of minimum Ct posteriors, 43 (7%) of proliferation duration posteriors, 64 (10%) of clearance duration posteriors, and 155 (25%) of peak timing ($t_p$) estimates had R values above 1.1. The $t_p$ parameter showed the highest frequency of poor convergence, and episodes with poorly estimated $t_p$ values almost always coincided with ill-fitting estimates of other individual-level parameters. These problematic $t_p$ fits were typically associated with infection episodes containing only a single positive sample, particularly when the Ct value was relatively high (indicating low viral load). Such weak or short-duration infection episodes frequently produced bimodal posterior distributions for $t_p$, reflecting greater uncertainty in peak timing. For type B influenza, it could also be the case that there is true biological variation as previous work has suggest that the viral shedding kinetics for type B influenza follow a highly variable and occasionally bimodal pattern [7,8].

Supplementary Fig. 2 visualizes a subset of the modeled viral shedding trajectories for five infection episodes for each subtype/lineage. These examples encompass the range of model performance, including two well-converged fits (R-hat = 1), one moderate fit (1.1 < R-hat < 1.5), and two poor fits (R-hat > 1.5). The poorly converged examples display distinctly bimodal, and in some cases trimodal, posterior estimates of the peak time parameter ($t_p$). Supplementary Fig. 3 presents the corresponding trace plots of the model posterior specifically for $t_p$, illustrating the sampling behavior underlying these convergence patterns. The bimodal and multimodal posterior distributions seen for these individuals arise primarily resulted from single-sample episodes, where insufficient temporal information constrained the model's ability to define a clear infection trajectory. It is also possible that in some cases, there was true biological variation; type B infections have previously been shown to exhibit bi- or multiphasic viral shedding kinetics [8].

Although such multimodality increases uncertainty in $t_p$ estimation and may reduce apparent model fit, the posterior mean typically falls between alternative peaks, providing a reasonable approximation of the overall infection course. The broader characteristics of the shedding profile, including total duration and magnitude, remain well represented, even in these uncertain cases. Small inaccuracies in peak timing could, however, introduce modest variability in downstream analyses that use $t_p$ to inform household-level force-of-infection (FOI) estimates. To incorporate this uncertainty, both the shedding characteristic regressions and the household transmission model regression analyses used 100 realizations of resampled posterior shedding trajectories across all subtypes, ensuring robust inference despite variable convergence quality.

#### 2.3 Visualizing the estimated shedding kinetics in the main text

In Figure 2A and 2E, when visualizing the average viral shedding kinetics for each subtype or strain, we first transformed the model-predicted $\Delta Ct$ values back to $Ct$ by adding the limit of detection threshold ($Ct_{lod}$) to enable direct comparison with the observed RT-PCR data. We then rescaled the temporal axis from calendar time ($t$) to time since peak shedding ($\tau$), defined relative to the average timing of minimum $Ct$ (i.e., the average timing of peak viral shedding, $\overline{t_P}$). These transformations are defined as follows:

$$Ct(\tau) = \Delta Ct(\tau) + Ct_{lod}$$

$$\tau = t - \overline{t_P}$$

#### 2.4 Regression analysis of risk factors associated with viral shedding duration and peak viral load

To examine factors associated with the total duration of viral shedding (defined as the sum of viral proliferation and clearance stages) and peak viral load, for each of the 100 realizations of the resampled posterior shedding trajectories, we used mixed-effects multivariable linear regression models.

The covariates included in the models were pre-season HAI titer (categorized as <1:40 or ≥1:40), age, sex, and HIV infection status. These variables were selected from a larger pool of potential covariates based on their improvement of model fit, as assessed by the Akaike Information Criterion (AIC), or their relevance to the study objectives (Table S2). To account for potential batch effects across study years, we included study year as a random effect in the regression model. This approach allows the model to estimate a separate intercept for each year while simultaneously estimating the variance of these intercepts across years. In doing so, the model captures unobserved year-to-year variability without assuming that such differences systematically influence the relationship being tested, for example due to differences in measurement or sample collection. For B/Yamagata, which circulated in only a single study year, inclusion of a random effect for year was not possible; therefore, a standard multivariable linear regression model was used instead. All reported p-values correspond to two-sided tests, with statistical significance defined as $p < 0.05$.

The specific equations of the regression models are as follows:

$$D_i = \beta_0^D + \beta_T^D x_i^T + \boldsymbol{\beta_A^D x_i^A} + \boldsymbol{\beta_S^D x_i^S} + \boldsymbol{\beta_{HI}^D x_i^{HIV}} + b_Y^D x_i^Y$$

$$P_i = \beta_0^P + \beta_T^P x_i^T + \boldsymbol{\beta_A^P x_i^A} + \boldsymbol{\beta_S^P x_i^S} + \boldsymbol{\beta_{HIV}^P x_i^{HIV}} + \boldsymbol{b_Y^P x_i^Y}$$

Where:

$D_i$ – shedding duration for infection episode $i$, scalar term.

$P_i$ – peak shedding intensity for infection episode $i$, scalar term.

$\beta_0^D$ – regression intercept for shedding duration, scalar term.

$\beta_0^P$ – regression intercept for peak shedding intensity, scalar term.

$\beta_T^D$ – regression coefficient for the association between pre-season HAI titer and shedding duration, scalar term.

$\beta_T^P$ – regression coefficient for the association between pre-season HAI titer and peak shedding intensity, scalar term.

$x_i^T$ – individual's pre-season HAI titer for infection episode $i$, scalar term.

$\boldsymbol{\beta_A^D}$ – regression coefficients for the association between age group and shedding duration, vector term, with each element of the vector representing the coefficient of a specific age group.

$\boldsymbol{\beta_A^P}$ – regression coefficients for the association between age group and peak shedding intensity, vector term, with each element of the vector representing the coefficient of a specific age group.

$\boldsymbol{x_i^A}$ – individual's age categories for infection episode $i$, vector term, with each element of the vector representing a specific age category.

$\boldsymbol{\beta_S^D}$ – regression coefficients for the association between sex and shedding duration, vector term, with each element of the vector representing the regression coefficient of a specific sex category.
$\boldsymbol{\beta_S^P}$ – regression coefficients for the association between sex and peak shedding intensity, vector term, with each element of the vector representing the regression coefficient of a specific sex category.
$\boldsymbol{x_i^S}$– individual's sex categories for infection episode $i$, vector term, with each element of the vector representing a specific sex category.

$\boldsymbol{\beta_{HIV}^D}$ – regression coefficients for the association between HIV status and shedding duration, vector term, with each element of the vector representing the regression coefficient of a specific HIV infection status.
$\boldsymbol{\beta_{HIV}^P}$ – regression coefficients for the association between HIV status and peak shedding intensity, vector term, with each element of the vector representing the regression coefficient of a specific HIV infection status.
$\boldsymbol{x_i^{HIV}}$– individual's HIV status categories for infection episode $i$, vector term, with each element of the vector representing a specific HIV status.

$\boldsymbol{b_Y^D}$ – random effects on shedding duration by calendar year, vector term, with each element of the vector representing the random effect of a specific calendar year.
$\boldsymbol{b_Y^P}$ – random effects on peak shedding intensity by calendar year, vector term, with each element of the vector representing the random effect of a specific calendar year.
$\boldsymbol{x_i^Y}$– calendar year when infection episode $i$ occurred, vector term, with each element of the vector representing a specific calendar.

Of the 623 infection episodes included in the viral shedding model, 50 samples tested positive for two influenza subtypes within the same specimen, corresponding to 25 co-infection events, and 77 reinfections were identified across study years. Most co-infections involved the two type A subtypes, which co-circulated during the first year of the study. Reinfections were most frequently observed between influenza A and influenza B viruses, typically when individuals were infected with influenza A early in the season and influenza B later in the same season. These co-infections and reinfections were retained in the shedding model to contribute to the population-level estimates for each subtype.

However, for the regression analyses assessing the association with pre-season HAI titer, co-infections and reinfections involving the same influenza type within a single season were excluded to avoid confounding due to immune interactions. Among the 77 reinfections, 21 involved the same subtype or type, resulting in the exclusion of 71 episodes in total (co-infections and reinfections combined). An additional 53 episodes were excluded because pre-season HAI titer measurements were unavailable due to missed samples, late enrollment, or sampling errors. The final dataset included 499 infection episodes, comprising 93 A(H1N1)pdm09, 149 A(H3N2), 66 B/Yamagata, and 191 B/Victoria infections.

Using the filtered dataset described above, we fitted regression models to each of the 100 realizations of the sampled viral shedding trajectories. Parameter estimates across realizations were subsequently pooled with the mice R package (version 3.17.0), thereby propagating uncertainty from the viral shedding kinetics into the final regression estimates. The pooled estimates are summarized in Fig 3 and Supplementary Fig 7. Linear and Poisson multivariable regressions on outcomes including shedding duration and peak shedding were conducted using package lme4 (version 1.1.34).

**S1 Supplementary Fig. 2. A sample of the estimated viral shedding kinetic fits from the shedding model.**

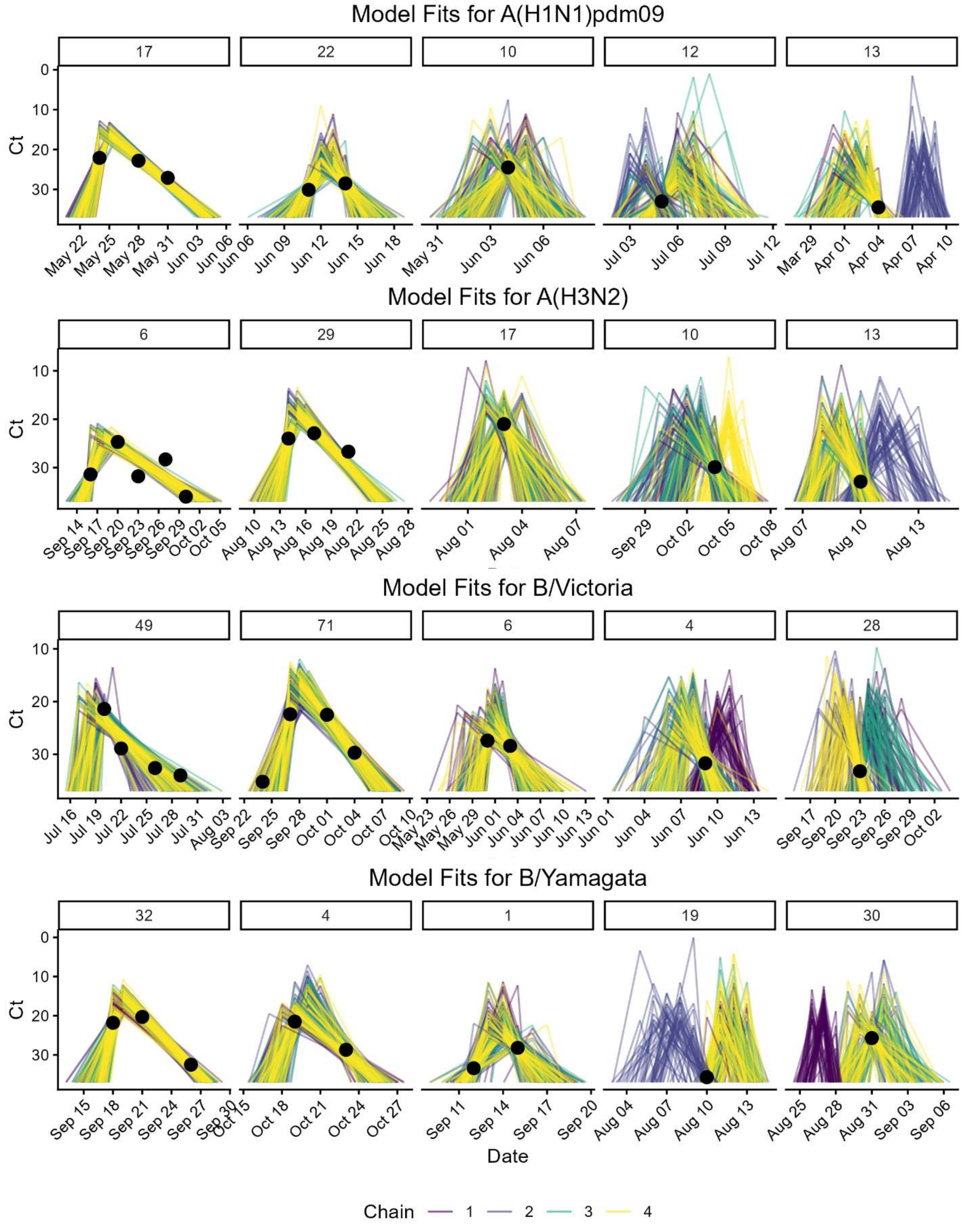

*Shown here are five infections for each subtype/lineage. Columns 1 and 2 present two strong fits (R-hat = 1), column 3 presents a one weak fit (1 < R-hat <1.1), and columns 4 and 5 present two poor fits (R-hat >1.1) . The dots are the raw Ct observations that informed the model; the colored lines are the first 50 iterations trace estimates of each chain from the model.*

**Supplementary Fig. 3. A sample of the model trace plots for estimation of the peak timing of viral shedding (tp).**

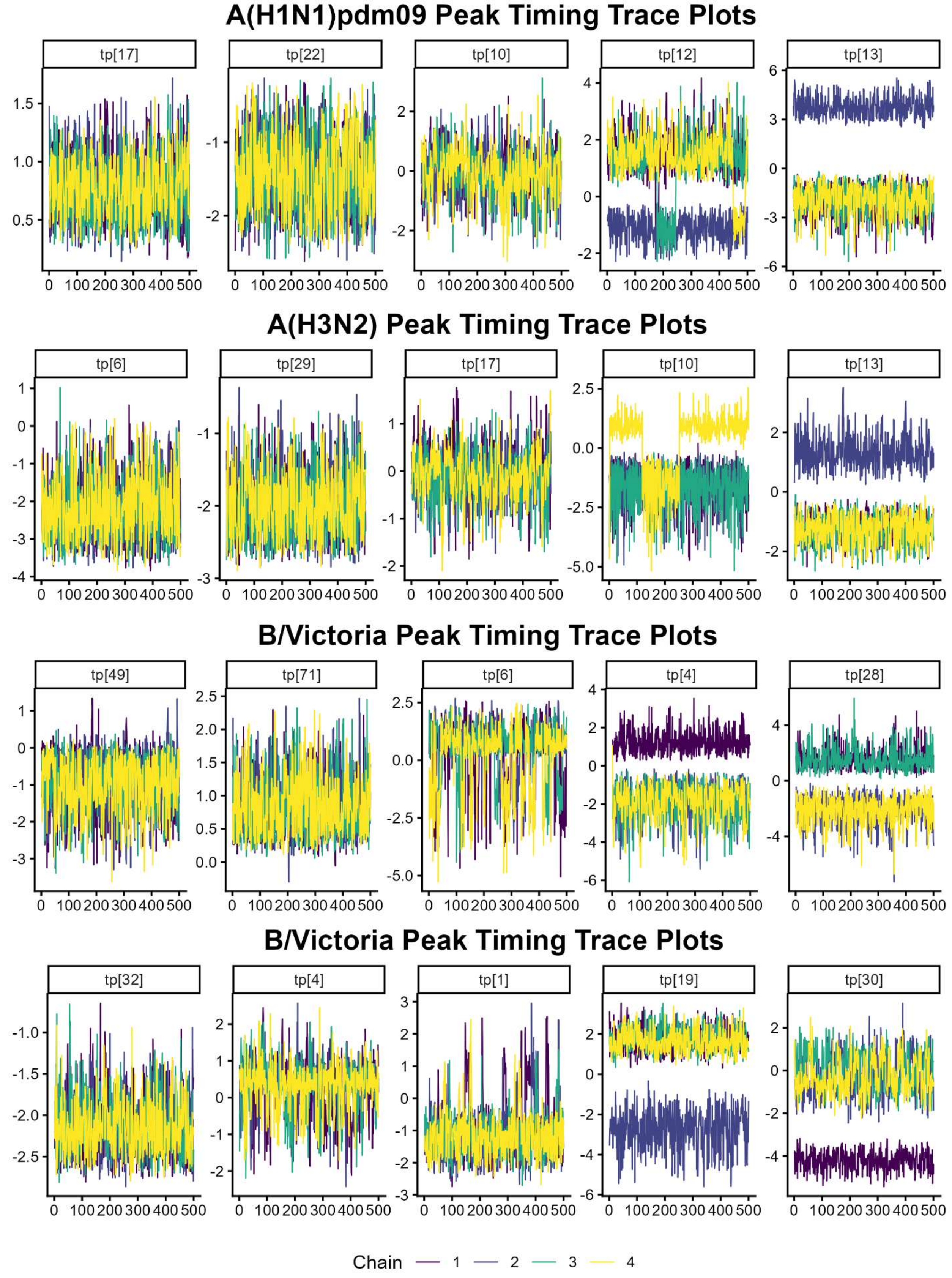

*Shown here are five infections for each subtype/lineage. The colored lines represent the estimates from the four different chains. Columns 1 and 2 present two strong fits (R-hat = 1), column 3 presents a one weak fit (1 < R-hat <1.1), and columns 4 and 5 present two poor fits (R-hat >1.1).*

### 2. Characterize risk factors associated with influenza household transmission

To characterize individual- and household-level risk factors associated with influenza transmission within households, we developed a regression-based transmission model that quantifies the probability of infection acquisition over time. This model links the risk of infection for each household member to both individual characteristics (e.g., age, pre-season immunity, and HIV status) and the household-level force of infection (FOI) derived from the viral shedding model, as well as the community-level risk of exposure.

Observations were created for each day in the influenza season, with a binary dependent variable indicating infection status. The estimated day of infection acquisition was derived from the shedding model, using the estimated duration of shedding and the day of peak infection: we subtracted the estimated proliferation duration from the peak day to obtain the first day of shedding, and, following prior literature, assumed infection acquisition occurred one day before shedding onset [9].

To improve the robustness of the household transmission model, the household and community forces of infection (FOIs) were informed by the characterized viral shedding trajectories of each influenza episode. The shedding model provided higher fidelity estimates of shedding onset compared to raw sampling data. Based on findings from human challenge studies[9,10], we assumed a one-day delay between the time of infection and the onset of viral RNA shedding. The binary dependent variable therefore reflected this estimated day of infection acquisition.

The household force of infection (FOI), defined as the sum of daily viral shedding intensities from infectious household members, was included as a time-varying covariate in the regression model. After accounting for household FOI, we further examined the effect of household size on infection acquisition to adjust for potential reductions in per-person contact intensity in larger households. This so-called "dilution effect," where an increase in household size is associated with a lower person-to-person transmission risk, has been reported in previous household transmission studies [11]. Including household size thus helps to contextualize household FOI in terms of exposure risk per contact, independent of household size.

The community FOI term was represented by the community-level prevalence of infection within the cohort at time t. This time-varying covariate was smoothed using a four-day moving average, corresponding to the maximum interval between consecutive sample collections. We used the cohort household data because households were randomly selected from the site populations, making cohort infection prevalence broadly representative of the community. Supplementary Fig. 10 shows time series of the smoothed community FOI, with separate figures for school-aged children (5–18) and other ages, with the school calendar indicated to provide context for outbreak timing. While school sessions could influence contact patterns, the community FOI shows no distinct timing differences, supporting its use as a reliable measure of population-level prevalence for the model.

Additional covariates included pre-season HAI titer (categorized by the 1:40 threshold), age, sex, HIV status, and household size (modeled as a continuous variable). The regression models were fitted separately for each influenza subtype to capture subtype-specific transmission dynamics.

Within the transmission model, we also excluded entire households in which one or more individuals had PCR-confirmed infection episodes but lacked estimable viral shedding trajectories, as the corresponding household FOI terms would otherwise omit a known source of infection exposure. The transmission model was implemented only for the season(s) in which a given influenza subtype was dominant; infections that occurred outside those dominant seasons were not included in the analysis. As in the shedding model, additional exclusions were made for individuals with missing pre-season HAI titer data.

The final dataset comprised 87 infection episodes, 923 individuals, and 206 households for A(H1N1)pdm09; 140 infection episodes, 500 individuals, and 105 households for A(H3N2); 165 infection episodes, 866 individuals, and 190 households for B/Victoria; and 64 infection episodes, 507 individuals, and 64 households for B/Yamagata.

To account for uncertainties in the viral shedding kinetics estimated in the previous section, we generated 100 realizations of the viral shedding trajectories by repeatedly resampling from the posterior distributions of the shedding model. For each of these 100 realizations and a given influenza subtype/lineage, a transmission model regression was fitted according to the following equation, which estimates the risk of infection acquisition for individual $i$ at calendar time $t$. This regression framework is equivalent to a discrete-time survival analysis of infection outcomes but allows the inclusion of time-varying covariates, such as community and household forces of infection (FOIs), without requiring the proportional hazards assumption [12–14]. The terms of the regressions are defined as follows:

$$y_i(t) = \text{logit}\left(\beta_C \cdot x^C(t) + \beta_V \cdot \sum_{j \neq i}^{j \in h}\left(\Delta Ct_j^L(t)\right) + \boldsymbol{\beta_T} \cdot \boldsymbol{x_i^T} + \boldsymbol{\beta_A} \cdot \boldsymbol{x_i^A} + \boldsymbol{\beta_S} \cdot \boldsymbol{x_i^S} + \boldsymbol{\beta_{HIV}} \cdot \boldsymbol{x_i^{HIV}} + \beta_{HS} \cdot x_i^{HS} + \alpha\right)$$

The terms are defined as follows:

$\beta_C$ – log of the hazard-ratio of infection associated with the community force of infection (FOI), scalar term.

$x_C$ – Community force of infection (FOI), represented based on the community-level prevalence of influenza infections at the study site at time t, scalar term.

$\beta_V$ – log of the hazard-ratio of infection associated with household FOI, scalar term.

$\sum_{j \neq i}^{j \in h}\left(\Delta Ct_j^L(t)\right)$ – household FOI at time t as the sum of the viral load of household members apart from the individual $i$ of interest, scalar term.

$\boldsymbol{\beta_T}$ – log of the hazard-ratio of infection associated with the pre-season HAI titer category (<1:40 or ≥1:40), vector term.

$\boldsymbol{x_i^T}$ – pre-season HAI titer category for individual $i$, vector term.

$\boldsymbol{\beta_A}$ – log of the hazard-ratio of infection associated with age group, vector term.

$\boldsymbol{x_i^A}$ – age group for individual $i$, vector term.

$\boldsymbol{\beta_S}$ – log of the hazard-ratio of infection associated with sex, vector term.

$\boldsymbol{x_i^S}$ – sex of individual $i$, vector term.

$\boldsymbol{\beta_{HIV}}$ – log of the hazard-ratio of infection associated with HIV status, vector term.

$\boldsymbol{x_i^{HIV}}$ – HIV status of individual $i$, vector term.

$\beta_{HS}$ – log of the hazard-ratio of infection associated with household size, scalar term.

$x_i^{HS}$ – household size of individual $i$, scalar term.

$\alpha$ – Poisson regression intercept, scalar term.

We fitted regression models to each of the 100 realizations of the sampled viral shedding trajectories. Parameter estimates across realizations were subsequently pooled with the mice R package (version 3.17.0), thereby propagating uncertainty from the viral shedding kinetics into the final regression estimates. The pooled estimates are summarized in Figures 5 and Supplementary

Fig 8. Linear and Poisson multivariable regressions on outcomes including shedding duration and peak shedding were conducted using lme4 (version 1.1.34).

### Supplementary Methods References

**Supplementary Fig. 4. Mosaic plot of nasopharyngeal swabs collected in 2016.**

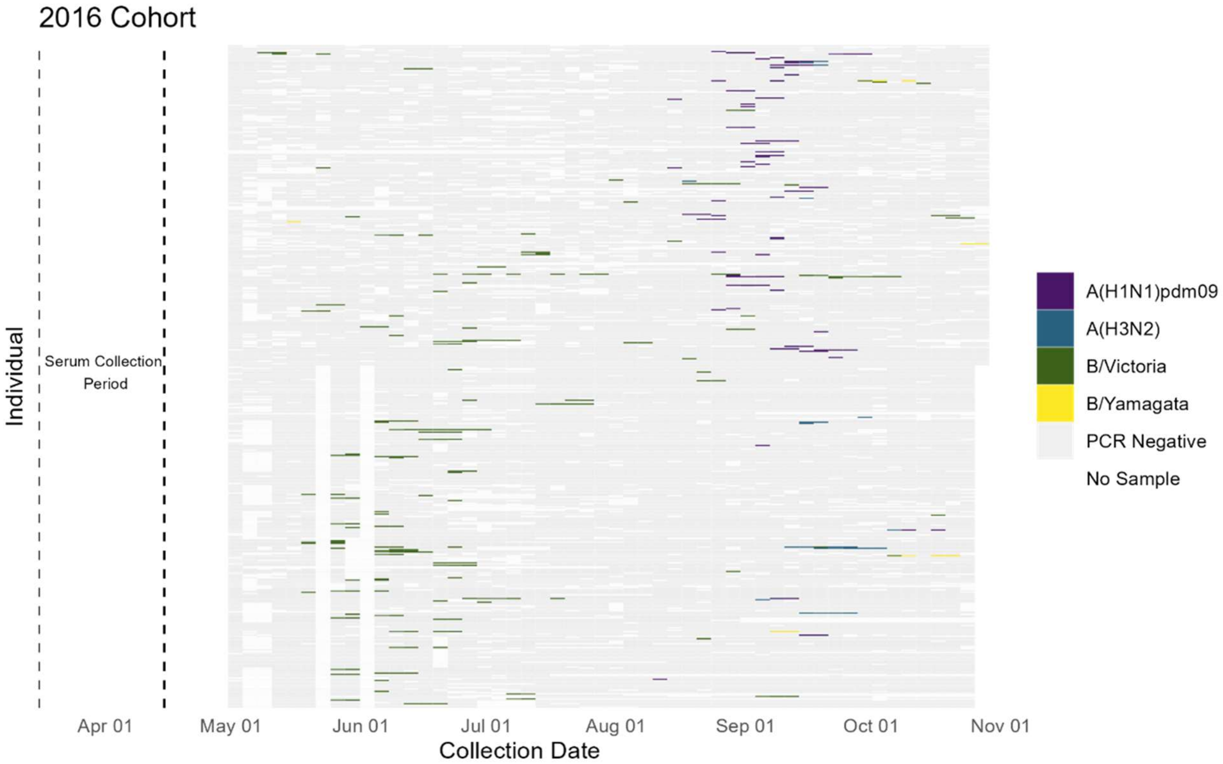


*Collection of nasopharyngeal swabs took place from May 5th through October 29th. The serum samples from which the HAI titers used in our analysis were collected between March 16th and April 15th. The color of the tile indicates the result of the swab, including the resulting subtype or lineage in the case that the swab was positive.*

**Supplementary Fig. 5. Mosaic plot of nasopharyngeal swabs collected in 2017.**

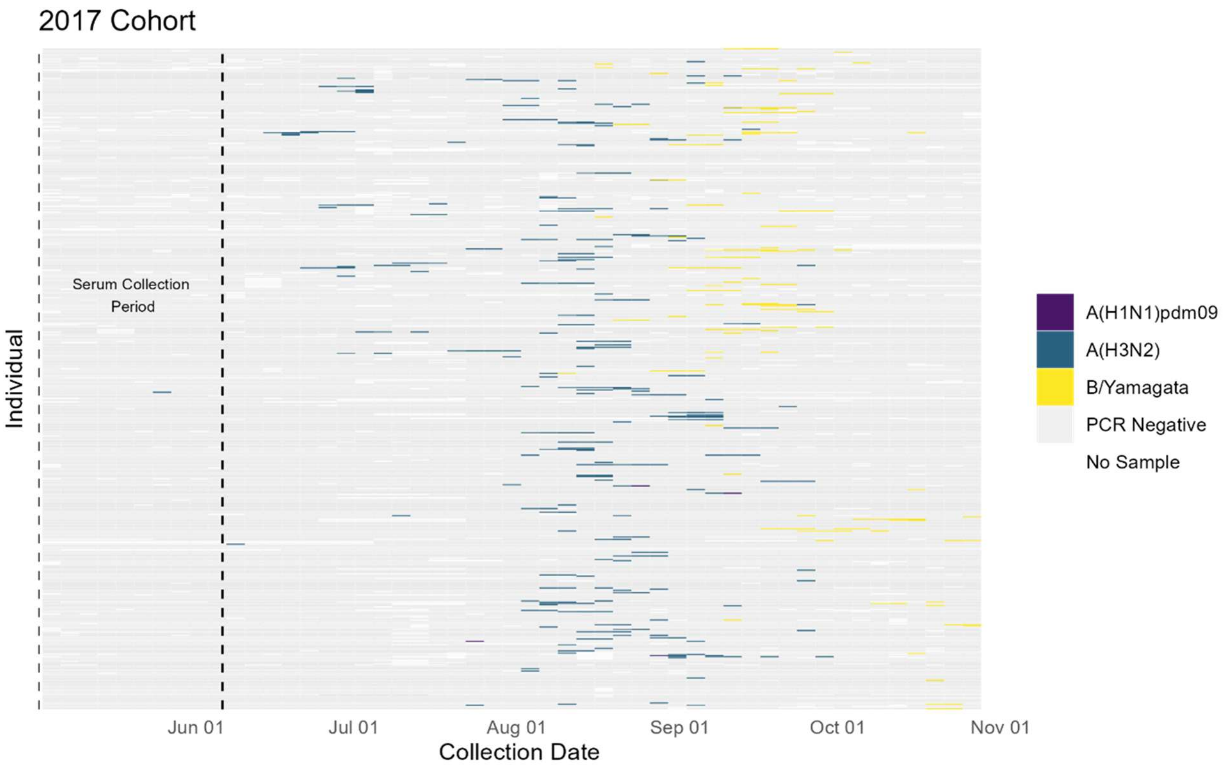


*Collection of nasopharyngeal swabs took place from January 16th through October 28th. The serum samples from which the HAI titers used in our analysis were collected between May 2nd and June 6th. The color of the tile indicates the result of the swab, including the resulting subtype or lineage in the case that the swab was positive.*

**Supplementary Fig. 6. Mosaic plot of nasopharyngeal swabs collected in 2017.**

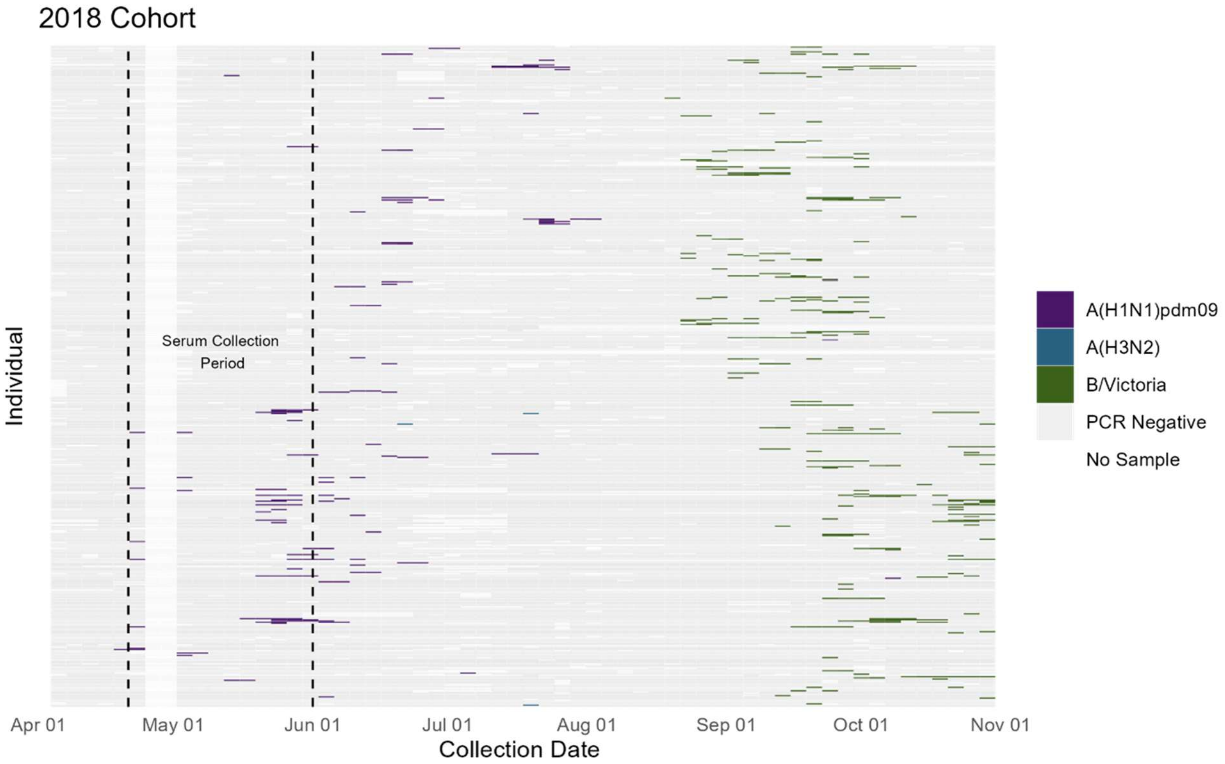


*Collection of nasopharyngeal swabs took place from January $15^{th}$ through October $31^{st}$. The serum samples from which the HAI titers used in our analysis were collected between April $21^{st}$ and June $1^{th}$; for individuals that had an infection during this period, a regression was used to estimate their titer for this period based on a sample from November – December of the previous year and the typical titer waning demonstrated in the cohort. The color of the tile indicates the result of the swab, including the resulting subtype or lineage in the case that the swab was positive.*

**Supplementary Fig. 7. Predictors of influenza virus shedding characteristics by subtype/lineage based on multivariable linear regression with HAI titer on a continuous log base 4 scale.**

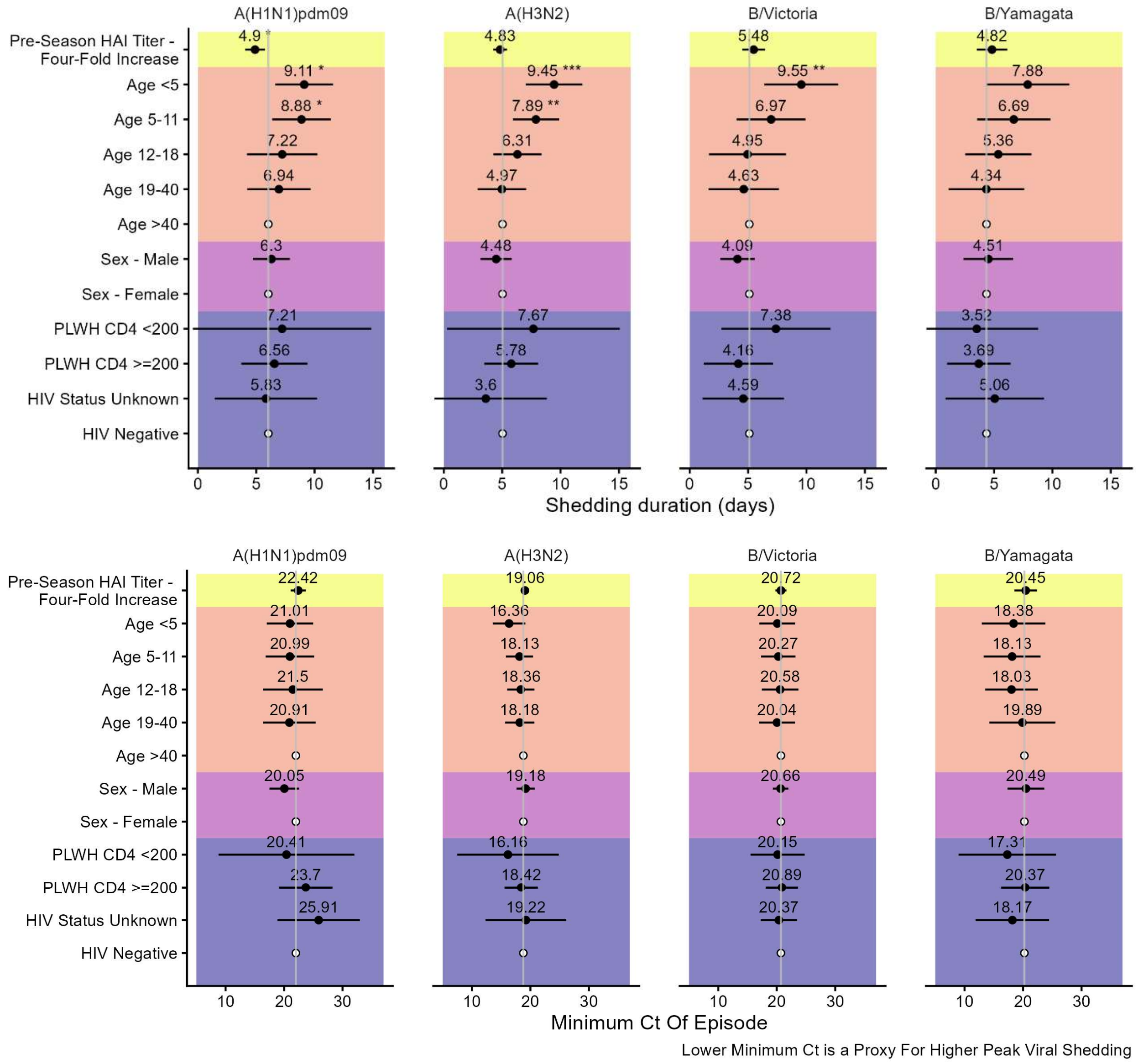


*The top panel regresses the total shedding duration and the bottom panel the minimum Ct of the infection episode, a proxy for the peak viral load. For the HIV status covariate, PLWH stands for person living with HIV further defined by demonstrating a CD4 T cell count below or equal to and above 200. Estimates are given in units of days with lines representing the 95% CI. The central*

*line indicates the mean duration for the subtype with the estimates given the unit of days. The reference groups are designated by a hollow dot. For the shedding duration the reference estimates for each subtype are as follows: A(H1N1)pdm09 4.26 days, A(H3N3) 4.77 days, B/Victoria 5.68 days, B/Yamagata 5.00 days. For the minimum Ct of episode the reference estimates are: A(H1N1)pdm09 22.60 Ct, A(H3N3) 19.24 Ct, B/Victoria 20.72 Ct, B/Yamagata 20.53 Ct. Asterisks denote p-value: ‘***’: <0.001 ‘**’: <0.01 ‘*’: <0.05 calculated via Satterthwaite's degrees of freedom method.*

**Supplementary Fig. 8. Factors influencing the risk of influenza virus infection acquisition by subtype/lineage based on multivariable logistic regression with HAI titer on a continuous log base 4 scale.**

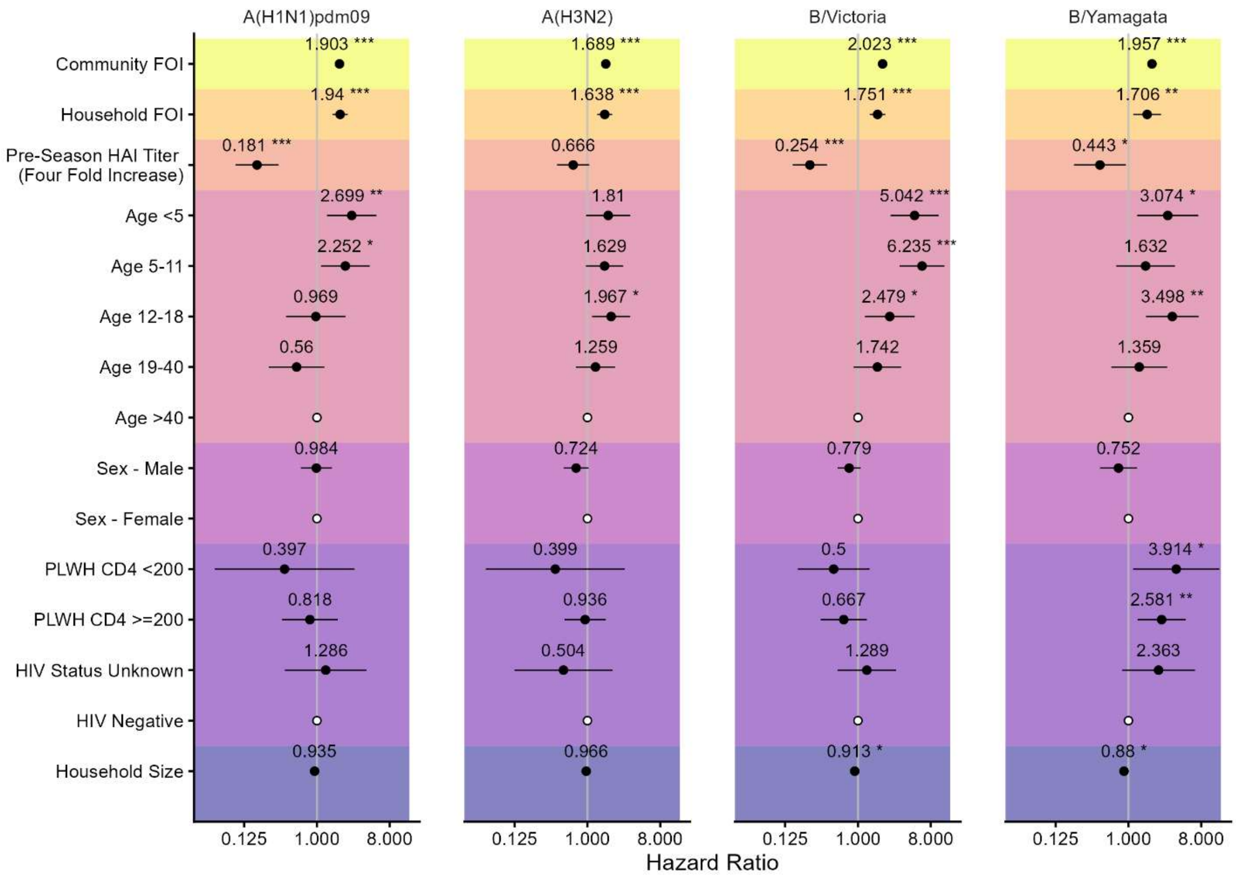


*Variables include the community force of infection (FOI), household FOI, pre-season HAI titer, age, sex, HIV status, and household size. Community FOI is indicated with respect to a 0.01 increase in absolute community prevalence (defined as the total number of infection episodes within the cohort at a given time / cohort population). Household FOI is indicated with respect to a step by 10 Ct units within the household. For the HIV status covariate, PLWH stands for persons living with HIV further defined by demonstrating a CD4 T cell count below or equal and above 200. Black dots represent point estimates of hazard ratios (logistic regression is equivalent to a Cox model in this context), with back horizontal lines indicating 95% CI intervals. The reference*

*groups for categorical variables are designated by a white dot. Asterisks denote p-value '***': <0.001 '**': <0.01 '*': <0.05.*

**Supplementary Fig. 9. Distribution of the pre-season HAI titers separated by observed infection in season.**

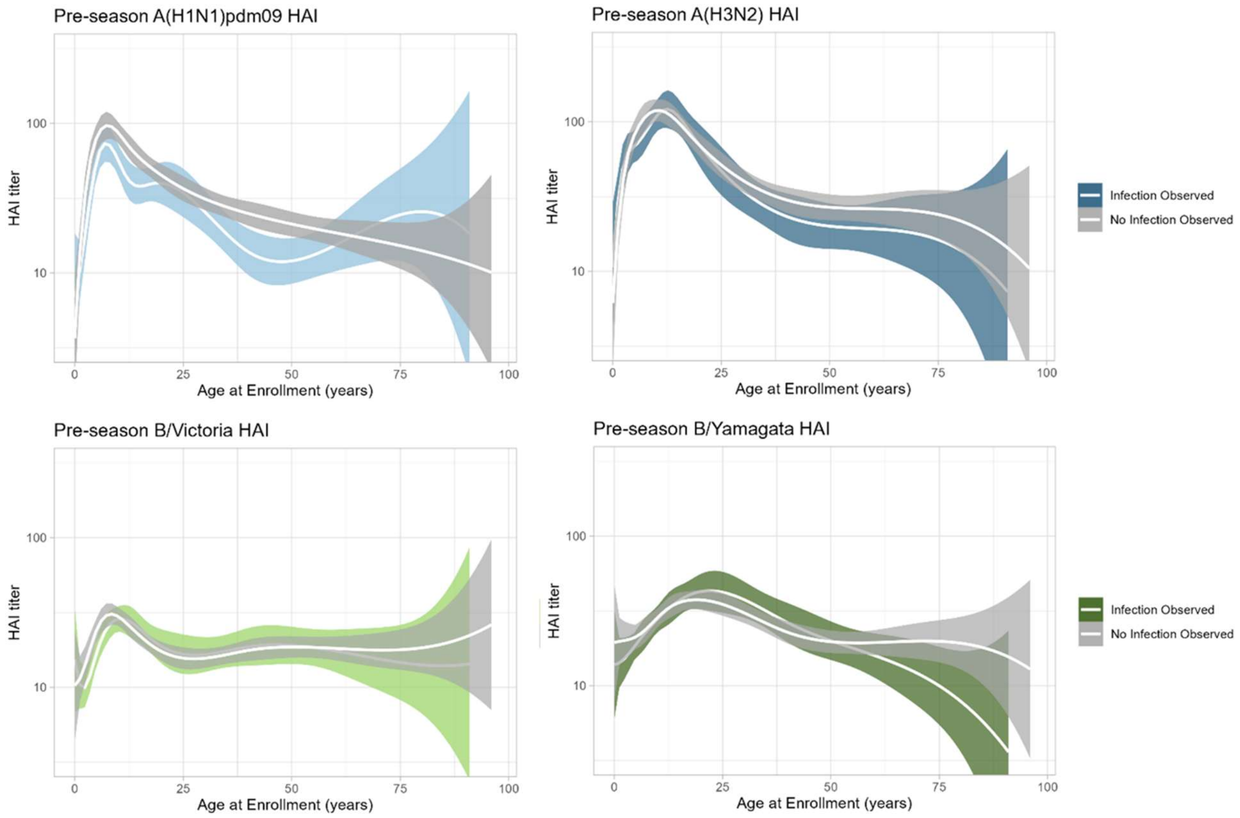


*The titer is given on a log base 10 scale and distributed by the age of the participant at enrollment, separated by subtype as indicated. The two lines indicate those who had an observed infection (colored shading) and those who did not have an observed infection (grey shading). The titers were tested with an assay against the recently circulating subtype. The white lines are a fitted spline linear spline fit with B-spline knots at ages 5, 12, 18, and 40 to describe age-dependent patterns, with the shaded region representing the 95% confidence interval.*

486 **Supplementary Fig. 10 Temporal trends in infection prevalence informing community FOI**

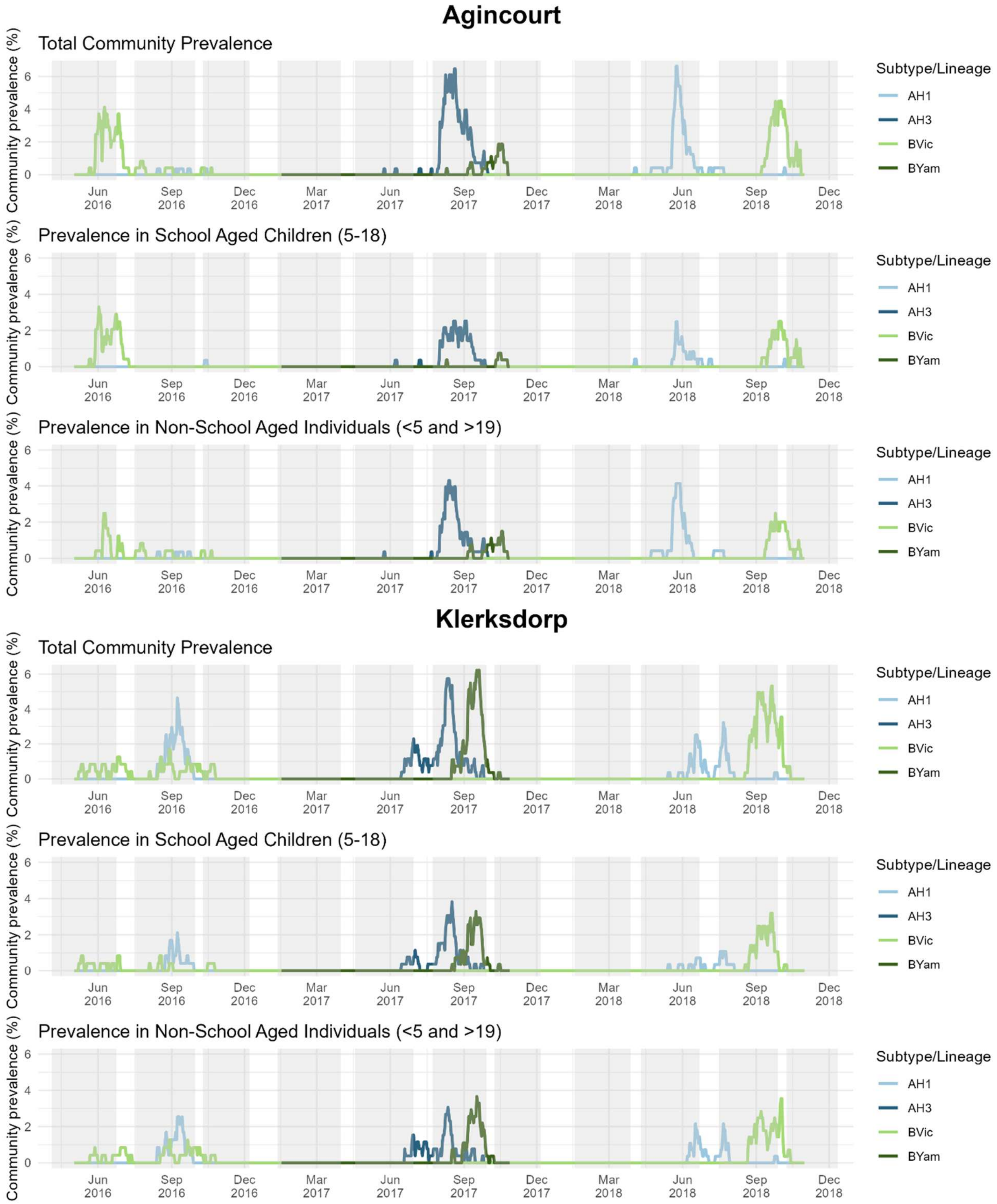


487 *Each set of three panels represents the two cohort sites, Agincourt and Klerksdorp. The top panels*

488 *of the sets show total community prevalence, calculated as the number of infections on a given day*

*divided by the total cohort population size and smoothed over a 4-day window to account for potential missing samples. The middle panels show prevalence restricted to school-aged individuals, and the bottom panels show prevalence among non-school-aged individuals. The colored lines correlate to the prevalence for each of the four subtypes/lineages. Community FOI, and thus the prevalence here, was only calculated for the years used in the transmission model for each subtype/lineage (2016 and 2018 for A(H1N1)pdm09 and B/Victoria, 2017 for A(H3N2) and B/Yamagata. The grey shaded regions in each figure indicate when school was in session.*

**Supplementary Fig. 11 Household cumulative infection risk by household size and influenza virus subtype/lineage**

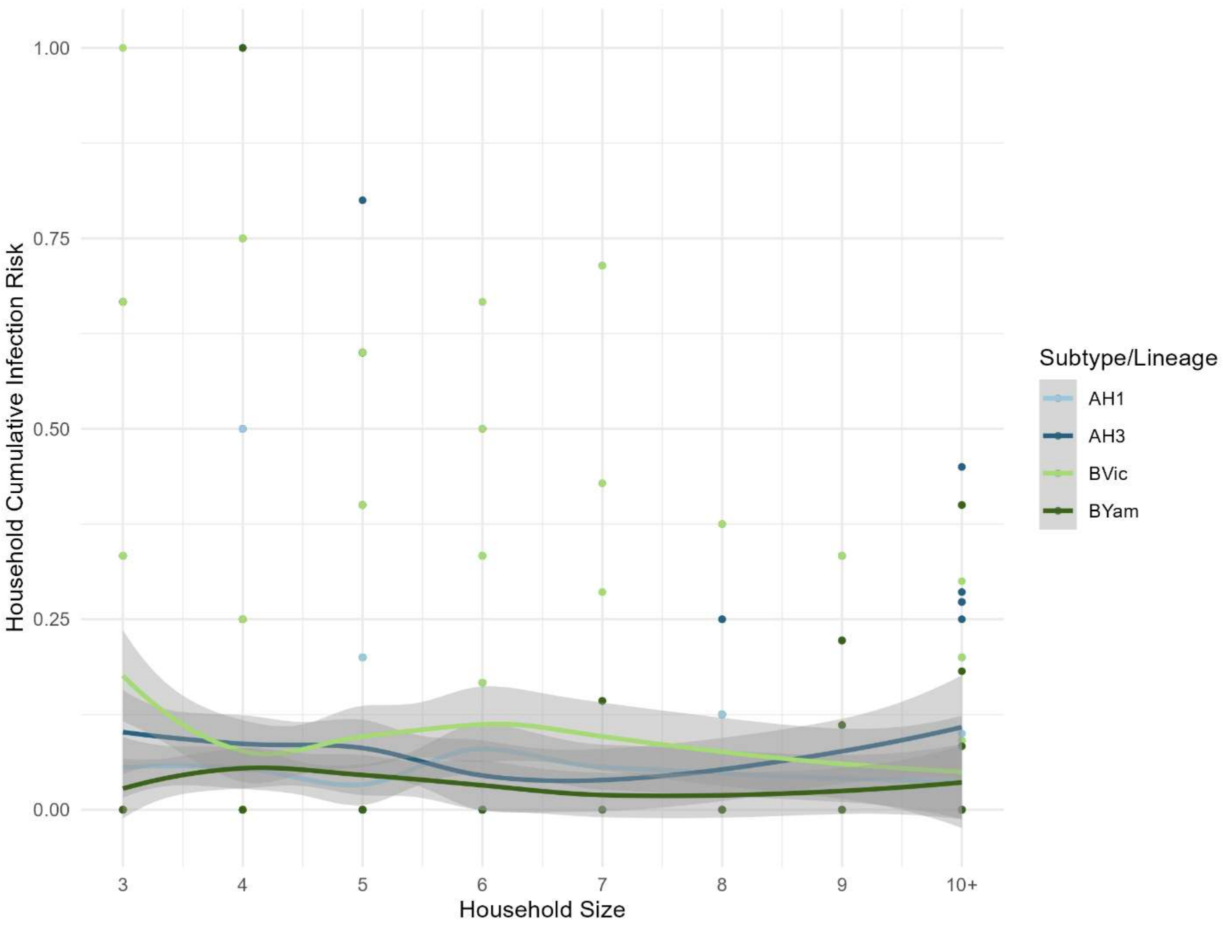


*Each point represents the observed cumulative infection risk for a given household size and influenza virus subtype or lineage. The cumulative infection risk was calculated as the proportion of individuals experiencing an infection within each household during the observed influenza season. Lines show locally smoothed trends in cumulative infection risk by household size. Household size was treated as a continuous variable from 1 to 10, with all households of size 10 or larger grouped into the "10+" category.*

**Supplementary Table 1. Coefficients of variation for the estimated proliferation and clearance shedding durations for each subtype/lineage.**

| Subtype/Lineage | Proliferation Coefficient of Variation | Clearance Coefficient of Variation |
|---|---|---|
| **A(H1N1)pdm09** | 28.0 | 82.6 |
| **A(H3N2)** | 69.3 | 78.3 |
| **B/Victoria** | 61.9 | 83.0 |
| **B/Yamagata** | 39.2 | 68.7 |

**Supplementary Table 2. Model building to determine variables included in the main model regression.**

| Subtype/ Lineage Model | Community FOI | + Household FOI | + Age | + Categorical (1:40) Pre-season Titer | + Sex | + HIV Status | + Household Size (**Main Model**) | + Year as fixed effect | + Year as random effect | Main Model + Study Site | Main Model + BMI | Main Model + Living With Child under 5 Years Old | Main Model + Crowding in Household |
|---|---|---|---|---|---|---|---|---|---|---|---|---|---|
| A(H1N1)pdm09 | 1501.3 | 1466.1 | 1445.2 | 1423 | 1425 | 1429.7 | 1430.7 | 1432.3 | Singular Fit | 1431.2 | 1436.2 | 1432.4 | 1432.5 |
| A(H3N2) | 1962.9 | 1944.9 | 1949.5 | 1946.8 | 1945.5 | 1948.6 | 1948.1 | | | 1946.7 | 1952.7 | 1950.1 | 1948.9 |
| B/Victoria | 2401.2 | 2381.9 | 2319.6 | 2309.6 | 2311.2 | 2315.2 | 2312.9 | 2299.8 | Singular Fit2306.3 | 2314.4 | 2319.4 | 2314.4 | 2314.8 |
| B/Yamagata | 959.82 | 958.2 | 961.15 | 961.23 | 959.85 | 954.94 | 950.68 | | | 951.69 | 956.14 | 950.98 | 951.97 |

*Values given represent the AIC for the described model. The highlighted cell represents the regression used for the main model. FOI stands for force of infection. BMI stands for body mass index.*

**Supplementary Table 3. Sensitivity analysis on the household force of infection (FOI) and community FOI terms included in the main model regression.**

| Subtype/Lineage Model | Main Model | Main Model Without Community FOI | Null Community Model (Binary Community FOI Term Representing Prevalent/Absent in Study Site) | Main Model Without Household FOI | Null Household Model (Binary Household FOI Term Representing Prevalent/Absent in Household) |
|---|---|---|---|---|---|
| A(H1N1)pdm09 | 1430.7 | 1439.1 | 1338.1 | 1473.2 | 1397.4 |
| A(H3N2) | 1948.1 | 2129.2 | 1901.6 | 1968.3 | 1945.4 |
| B/Victoria | 2306.3 | 2532.2 | 2286.7 | 2338 | 2285.4 |
| B/Yamagata | 950.68 | 1082.9 | 922.03 | 952.03 | 946.97 |

*Values given represent the AIC of the given model. For the community null model, even though the model fits better, the estimates become very high (ex. 42 odds ratio or a 4000% increase in risk of H3 infection acquisition, which suggests a vast oversimplification of the parameter). Similarly, though the null household model fits slightly better for the null household model, in the A(H1N1)pdm09 regression, the community FOI is no longer significant meaning that there would be no significant risk of infection acquisition from community exposure suggesting again an oversimplification of the parameter that confounds the results.*
